# Characterization of cell states in biliary tract cancers identifies mechanisms of therapeutic resistance in a phase II trial of DKN-01/nivolumab

**DOI:** 10.1101/2024.10.08.24315092

**Authors:** Ryan J Park, Milan Parikh, Leon Pappas, Moshe Sade-Feldman, Anupriya S. Kulkarni, Lynn Bi, Thomas J. LaSalle, Aralee Galway, Caroline Kuhlman, Lawrence S Blaszkowsky, Jeffrey A. Meyerhardt, Peter C Enzinger, Leah Biller, Jill N Allen, Michael H. Kagey, Jason Baum, Cynthia Sirard, Dan G. Duda, Andrew X. Zhu, Thomas A. Abrams, Nir Hacohen, David T. Ting, Arnav Mehta, Lipika Goyal

## Abstract

Biliary tract cancers demonstrate profound therapeutic resistance, and broadly effective therapies for refractory disease are lacking. We conducted a single-arm, second-line phase II trial combining DKN-01, a humanized monoclonal antibody targeting Dickkopf-1 (DKK-1), and nivolumab to treat patients with advanced biliary tract cancer (NCT04057365). No objective responses were seen. To identify mechanisms of treatment failure, we analyzed paired pre-treatment and on-treatment biopsies using scRNA-seq and constructed a detailed molecular classification of malignant and immune cells. We annotated five biliary tract cancer malignant cell states: classical, basal, mesenchymal, neural-like, and endothelial-like. Neural-like and endothelial-like states, which drive therapeutic resistance in other cancers, have not previously been described in BTC. Malignant cell states co-varied with distinct immune cell states, revealing diverse mechanisms of myeloid and T-cell mediated immune suppression, including M2 myeloid and terminally exhausted T cell programs that were induced by DKN-01/nivolumab. Here, we provide the first systematic classification of functionally annotated cell states in biliary tract cancer and provide new insight into resistance mechanisms to an immunotherapy combination that can inform the next generation of trials.

## Introduction

Biliary tract cancers (BTCs) are rare and agressive malignancies of the bile ducts and gallbladder with a 5-year overall survival (OS) rate ranging between 2-30% depending on the disease stage at diagnosis^1,2^. For over a decade, standard first-line therapy for patients with unresectable or metastatic BTC has consisted of combination chemotherapy with gemcitabine and cisplatin^3^. Recently, two phase 3 trials testing the addition of durvalumab^4^ or pembrolizumab^5^ to this chemotherapy backbone demonstrated a survival improvement, offering a median overall survival of approximately 13 months and offering durable benefit in approximately 15% of patients^4,6^. These studies highlighted for the first time the potential of immunotherapy combination strategies in unselected patients with BTC^7,8^. Despite successes, median OS was prolonged by less than 2 months, reflecting the fact that most BTCs have primary resistance or rapidly adapt resistance to immunotherapy. A major obstacle to progress is that neither tumor cell states nor the immune composition of the BTC tumor microenvironment (TME) are well characterized^7,8^.

Molecular subtyping using both genomics or transcriptomics of cancer cells has been successfully developed in lung^9–11^, pancreatic^12,13^, melanoma^14,15^, breast^16^, and prostate cancer^17^, among others, and lays the foundation for understanding the heterogeneity of tumor states and within the TME. However, no consensus classification of BTC subtypes currently exists^18,19^. BTC is anatomically classified as intrahepatic, perihilar, and distal cholangiocarcinoma and gallbladder cancer. Multiple histologic subtypes within this anatomic classification correlate with distinct molecular profiles, suggesting potentially different cells of origin^20^. Recent bulk and single-cell RNA-sequencing (scRNAseq) analyses have demonstrated transcriptionally and cellularly distinct subtypes of intrahepatic cholangiocarcinoma that underlie both intratumoral and intertumoral heterogeneity^19,21–23^. Moreover, prior scRNAseq and single-cell ATAC-seq (scATACseq) studies in intrahepatic^24,25^ and distal cholangiocarcinoma^26^ have shown diversity in the TME cellular composition between patients. Importantly, the implications of differential immune infiltration on immunotherapy treatment response and survival has been more recently characterized^18,27^. Notably, these studies did not subtype malignant cells reproducibly across patients, including in other tumors of similar developmental origin. Furthermore, they did not define tumor cell states, a practice observed in other tumors through gene programs learned from tumor cells^28,29^. This characterization has implications for understanding plasticity and therapeutic resistance^30^.

Other than the recent limited additive activity of durvalumab or pembrolizumab to first-line chemotherapy backbone therapy, immune checkpoint inhibitors as monotherapy has been ineffective for BTC with objective response rates of less than 10% for patients with microsatellite-stable (MSS) tumors, and up to 22% in a trial for which MSS status was not reported^31,32^. Limited responses to immune checkpoint inhibitors are hypothesized to be due to the: (a) tolerogenic environment of the liver^33^, (b) desmoplastic environment of the tumor^34^, (c) heterogeneous tumor phenotypes with varying immune subsets^35,36^, and (d) several cellular TME immunosuppressive mechanisms^34,37,38^. These hypotheses are supported by a scRNAseq study of baseline and anti-PD1-treated cholangiocarcinoma biopsies^36^.

One relatively new molecular target in epithelial cancers is dickkopf-related protein 1 (DKK1), a modulator of Wnt signaling pathways and an inhibitor of the canonical Wnt/β-catenin signaling pathway^39^, with pleiotropic activities in other pathways. Wnt/beta-catenin signaling has been shown to influence several important processes, such as cell growth and differentiation, and bone development^40–44^. DKN-01 is a humanized IgG4 monoclonal antibody that antagonizes the DKK1-LRP6 interaction. In a study of 138 intrahepatic cholangiocarcinoma samples, 38% of intrahepatic cholangiocarcinoma samples demonstrated DKK1 expression, and high expression was associated with lymph node metastasis, advanced tumor stage, and lower 5-year survival rates^45^. Importantly, DKK1 also has immunomodulatory activity^39,46–48^ and has been shown to increase myeloid-derived suppressor cell (MDSC) infiltration^46–50^ and enhance the presence of M2-type macrophages that inhibit CD8 T and NK cell function. Given the possible complementary immune-stimulating effects, the combination of DKN-01 and immune checkpoint inhibitors is under investigation in several tumors^51^ (NCT04363801, NCT04166721, NCT05761951).

We conducted a phase II study to assess the synergistic role of DKN-01 and PD-1 blockade with nivolumab in patients with advanced BTCs who had tumor progression on at least one prior line of systemic therapy. We obtained paired biopsies both at baseline and 3 weeks after initiation of trial therapy and performed scRNAseq, characterizing novel molecular subtypes of BTC, distinct immune TMEs associated with each subtype, and the effects of the trial therapy on the TME. In this report, we present a novel framework for molecular subtyping of BTC, which is reproducible across patient samples, and we determine distinct tumor immune infiltrates that correlate with different tumor subtypes and lack of response to immunotherapy in BTC.

## Results

### Safety and efficacy of clinical trial

Between October 2019 and August 2020, 13 patients with advanced BTC were enrolled and treated across 2 sites. Each patient received DKN-01 600mg intravenously and nivolumab 240mg intravenously every 2 weeks until treatment progression or intolerance. The median age of patients at the time of consent was 66 years (range 53-77 years) (**Table S1**). All patients were white (100%) and the majority were female (61%) (**Table S1**). All thirteen patients had metastatic disease (100%), with the prevailing BTC type being intrahepatic cholangiocarcinoma (54%) and the remaining having extrahepatic cholangiocarcinoma (39%) and gallbladder adenocarcinoma (8%) (**Table S1**). All patients had an ECOG status of 0 or 1 and had received prior systemic chemotherapy but no prior DKK1 inhibitor or anti-PD-1/PD-L1 treatment, as per the eligibility criteria for the study (**Table S1**).

The combination of DKN-01 and nivolumab was overall well tolerated. The most common treatment-emergent adverse events included AST elevation (69%), anemia (54%), dyspnea (54%), hyponatremia (54%), ALT elevation (46%), and fatigue (46%) (**Table S2**). The most frequent grade 3 or 4 treatment-emergent adverse events included abdominal pain (15%), lymphopenia (15%), ALT elevation (8%), AST elevation (8%), alkaline phosphatase elevation (8%), dyspnea (8%) and hypokalemia (8%) (**Table S2**). One patient had a Grade 3 or 4 adverse event related to DKN-01 (lymphopenia) and one person died on protocol after recurrent admissions for bowel obstructions attributed to her malignancy after a single dose of study drugs.

The median treatment duration on protocol was 50 days (range 28 to 231 days). Among the 13 patients who received at least one cycle of DKN-01 and nivolumab, no objective responses were seen (0% ORR). Three patients had a best overall response (BOR) of stable disease (SD, 23%), 7 patients had a BOR of progressive disease (PD, 54%), 2 patients were deemed to have clinical progression (15%), and 1 patient passed away on protocol (8%) **(Table S3)**. The median PFS was 49 days (range 28 to 221 days, 95% CI 40-96 days).

### Remodeling of the tumor microenvironment after DKN-01 and nivolumab

To assess the effect of combined DKN-01 and nivolumab on the TME of BTC, we obtained 11 biopsies from 8 patients, which included 8 pre-treatment biopsies and 3 paired on-treatment biopsies taken at approximately 3-weeks after treatment start (**Table S3**). We performed scRNAseq of a total of 49,380 high-quality cells obtained from fresh biopsies (**Figure 1A**). We obtained representation of all major cell subsets expected in the TME and annotated tumor cells using marker genes and inferred copy number changes from transcriptional profiles (**Figures 1B,C and S1**). The abundance of cancer cells in each sample was variable, between 0.5% and 88.5% in pre-treatment samples (**Figure S2**). Of note, by scRNAseq, DKK1 expression was limited to 7 out of 11 patient samples and 883 out of 49,380 total cells (0.018%) (**Figure S3A-C**). 91% of all DKK1+ cells were tumor cells and the majority of the remainder were myeloid cells (**Figure S3D,E)**. In-situ hybridization (ISH) for DKK1 expression^52^ separately showed absence of DKK1 staining in 5 of 8 samples (**Figure S3B**), though notably this subset of samples largely did not overlap with the scRNAseq cohort. Overall, DKK1 expression was very low in all the patients enrolled on this trial and was not included in eligibility criteria as it was not a validated biomarker for DKN-01 efficacy at the time of study design. In addition, negative or 1+ PD-L1 staining was observed in 7 of 8 samples (**Figure S3C**).

**Figure 1.**
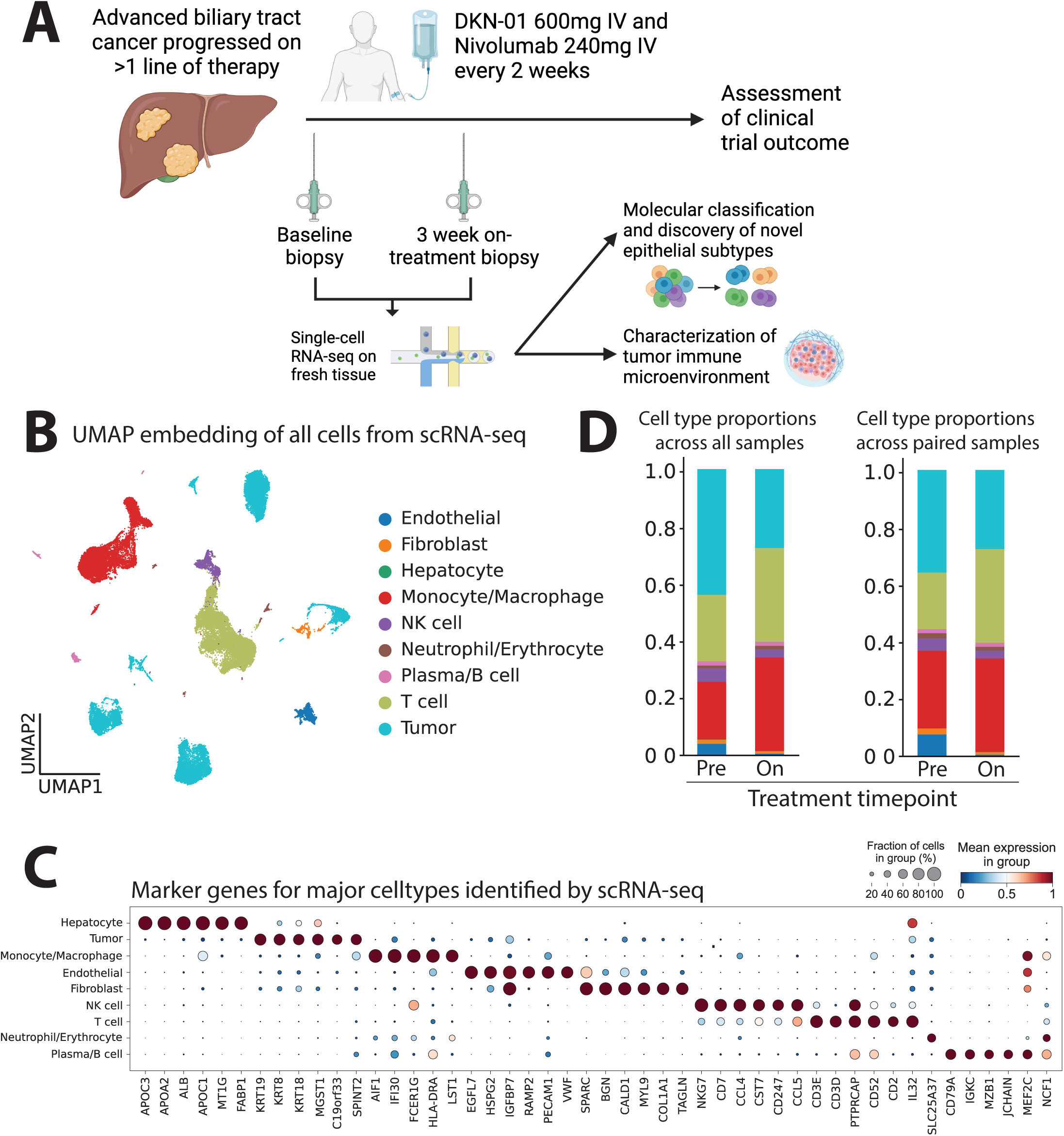
Single-cell profiling of paired patient biopsies in a clinical trial of biliary tract cancers patients treated with DKN-01 and nivolumab. (A) Schematic of trial treatments, timing of biopsies and analysis performed in this study. (B) UMAP embedding of single-cell RNA-seq (scRNAseq) profiles of all cells obtained across biopsy samples labeled by major cell types. (C) Dot plot showing expression level and proportion of cells expressing marker genes for all major cell types from scRNAseq data. Differential expression analysis was performed using a Wilcoxon rank-sum test. (D) Cell type proportions of all major cell types in baseline and on-treatment samples analyzed using all samples (left; n=11) and only-paired samples (right, n=6).

An analysis of the differential abundance of cell subsets (**Figure 1D**) revealed contraction of the tumor cell fraction and expansion of the T cell fraction in paired on-treatment biopsies relative to pretreatment biopsies (n=3 patients), though these changes were not statistically significant. Despite the low numbers of DKK1 positive cells, we found that the fraction of DKK1 positive tumor cells was significantly higher in pre-treatment samples compared to paired on-treatment samples (p=0.028, student t-test). Analysis of gene expression programs for Wnt signaling in addition to other components of the Wnt pathway, such as the Wnt co-receptor low-density lipoprotein receptor-related protein 6 (LRP6) and the Frizzled receptors (FZD), revealed heterogeneous expression beyond cancer cells, i.e., in endothelial cells, myeloid cells, T cells, erythroblasts, and fibroblasts (**Figure S3D-F**). These expression patterns suggest that the DKN-01-mediated blockade of the DKK1-LRP6 interaction may affect diverse cell types within the TME of BTC.

### Characterization of treatment-related changes in immune populations

Given the expansion of the T cell fraction observed in post-treatment samples, we next sought to further characterize the changes in the immune TME after treatment. We performed granular annotations of T cell, myeloid, and NK cell subsets based on marker gene expression (**Figures 2A-E and S4**). We examined the expression of a Wnt signaling gene set and CKAP4 (alternative receptor for DKK1) in these cell subsets and found the highest expression of Wnt signaling in naive/memory CD4+ T cells, resident memory, effector memory, and exhausted CD8+ T cells, and macrophages, whereas highest expression of CKAP4 was in fibroblasts and monocytes (**Figures S5 and S6**).

**Figure 2.**
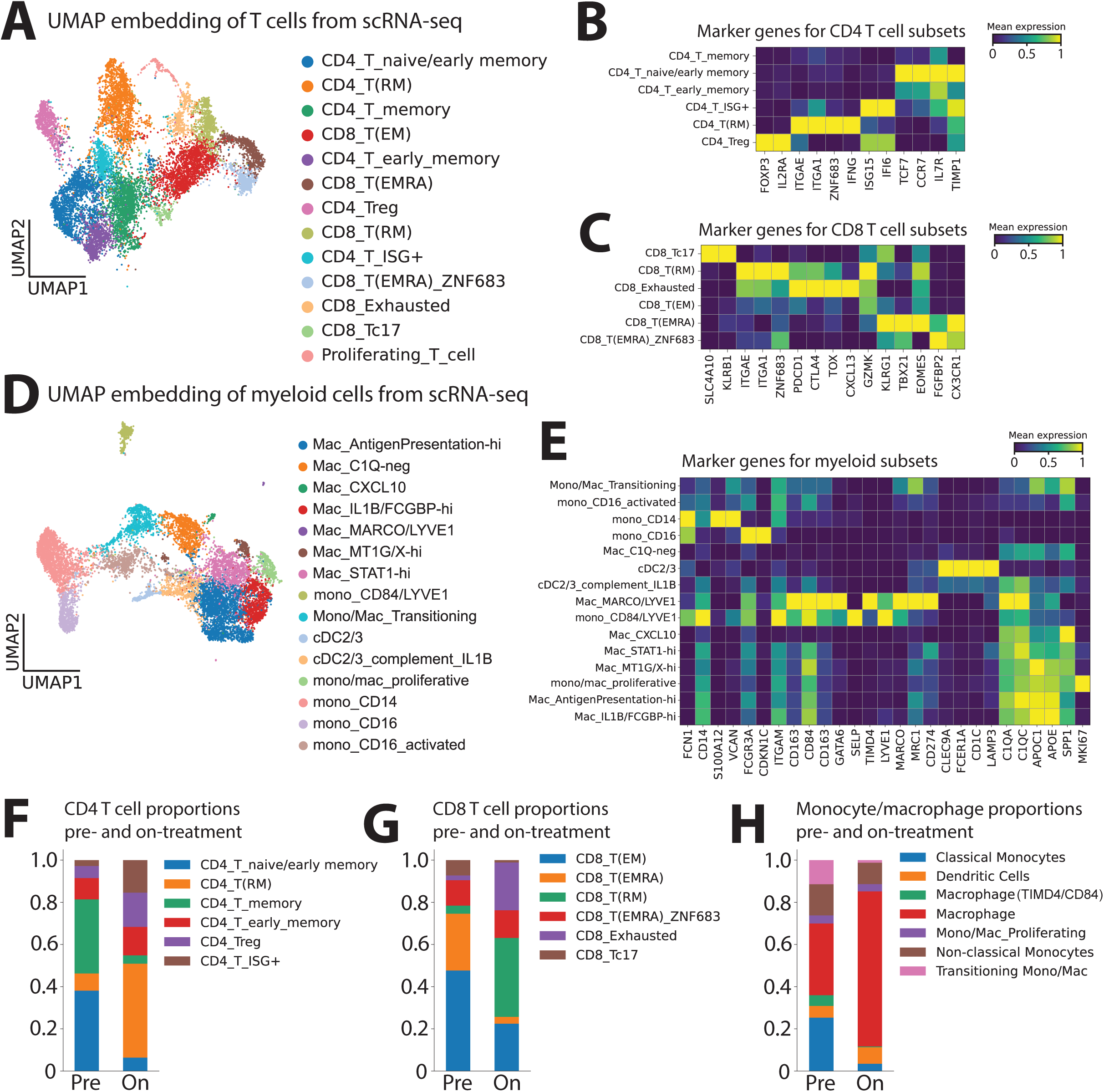
DKN-01 and nivolumab treatment leads to a redistribution of immune cell types in the tumor microenvironment. (A) UMAP embedding of all T cells profiled by scRNAseq labeled by granular subsets. Heatmap showing marker genes for (B) CD4+ and (C) CD8+ T cell subsets. (D) UMAP embedding of myeloid cells profiled by scRNAseq labeled by granular subsets. (E) Heatmap showing marker genes for myeloid cell subsets. Proportion of granular (F) CD4+ T cell, (G) CD8+ T cell and (H) myeloid cell subsets relative to their parent group in pre- and on-treatment biopsies.

Next, we examined the trends seen with DKN-01 and nivolumab treatment on redistributing immune cell populations in the TME. We found that in the CD4+ T cell compartment, there was a notable expansion of ISG+ and resident memory cells, whereas there was a contraction in memory and naive/early memory cells (**Figure 2F**). In the CD8+ T cell compartment, there was expansion of exhausted and resident memory cells, and contraction of effector memory cells (**Figure 2G**). The observed changes are consistent with the known effects of anti-PD1 treatment on tumor T-cell subsets. In the myeloid compartment, we observed a contraction of monocyte subsets and an expansion of macrophages (**Figure 2H**). We also identified a population of myeloid cells marked by expression of *CD84*^53^ (which regulates immunosuppressive activity and MDSC-like function), *CD163* (a canonical marker of immunosuppressive macrophages), and *TIMD4*^54,55^ (which marks mature, self-renewing tissue-resident and peritoneal macrophages) which were decreased in on-treatment samples. These data, therefore, are suggestive of rearrangement of the T cell and myeloid compartments in the TME of BTCs with DKN-01 and nivolumab despite the absence of clinical response.

To better understand the gene expression programs that are altered with treatment, we next performed consensus non-negative matrix factorization (cNMF) on the expression matrices of all T cells (**Figure S7,8 and Table S4**). We discovered eight gene programs in T cells, several of which were cell state-specific and others more globally expressed (**Figure S7A**). To characterize the programs in more detail, we scored all T cells on gene expression programs identified in T cells from a recent colorectal cancer (CRC) atlas^29^, and we looked at the correlation of expression of these programs with those identified in our dataset (**Figure S7B**). Among the eight BTC T cell programs we identified, we found ones highly correlated with programs for translation, GZMK, proliferation, and Tregs and CXCL13+ cells. Of note, we found increased expression of the GZMK program after treatment in the CD4+ ISG subset and decreased expression of the same program after treatment in CD8+ effector memory and exhausted subsets (**FIgure S7C**). Additionally, we found increased expression of the CXCL13 program in CD4+ resident memory and CD8+ exhausted subsets, which may be consistent with the expansion of tumor-reactive CD8+ T cells (**Figure S7C**).

Using the same approach as for T cells, we identified 14 gene expression programs in myeloid cells, which analogously demonstrated a mix of myeloid cell subtype-specific and global expression (**Figure S8A**). Correlating program expression with those from the CRC atlas enabled clear identification of programs related to S100 and C1Q subsets, glycolysis, ISGs, and proliferation (**Figure S8B**). Within high level myeloid subsets, we saw strong induction of the ISG program with treatment in non-classical monocytes, and weaker induction in macrophages (**Figure S8C**).

### Molecular subtyping of tumor cells in BTC

We next studied tumor cell states in our samples given the growing body of evidence to suggest that cell state plays a role in primary and adaptive resistance^14,28,30,56,57^. We first performed unbiased clustering of all tumor cells and found that cells clustered largely by tumor histology and metastatic tissue sites (**Figure S9A**). Recently, two subsets of intrahepatic cholangiocarcinoma have been described^58^. In our data, SPP1 was strongly expressed in intrahepatic cholangiocarcinoma and S100P had predominant expression in distal bile duct and gallbladder adenocarcinoma (**Figure S9A**), but there were also many cells without either SPP1 and S100P expression. Given that there are few well-described gene sets that have been used to molecularly characterize BTC tumors, we performed cNMF on the expression matrices of all tumor cells to identify forty-four gene programs within our dataset (**Figure S9B**) and also scored all tumor cells using gene programs derived from pancreatic adenocarcinoma (PDAC) which is anatomically closely related to biliary tract cancers^28,59^. We identified five distinct tumor cell states using these gene programs: (1) classical, (2) basal-like, (3) mesenchymal, (4) neuronal-like, and (5) a novel, uncharacterized state (**Figures 3A, S10A and Table S5**). Basal cells had strong expression of keratins (KRT7, KRT19, and KRT6A), mesenchymal cells expressed collagens (COL3A1, COL6A2, COL1A1) and several known mesenchymal markers (SPARC, VIM), neural cells expressed genes implicated in neural development (GLIS3, ZBTB20) and classical cells expressed several characteristic genes of epithelial states (TSPAN8, TFF1, EPCAM and CDH1) and S100P (**Figure 3B**). Tumor cells with activity of both mesenchymal and neural-like gene programs were annotated as intermediate.

**Figure 3.**
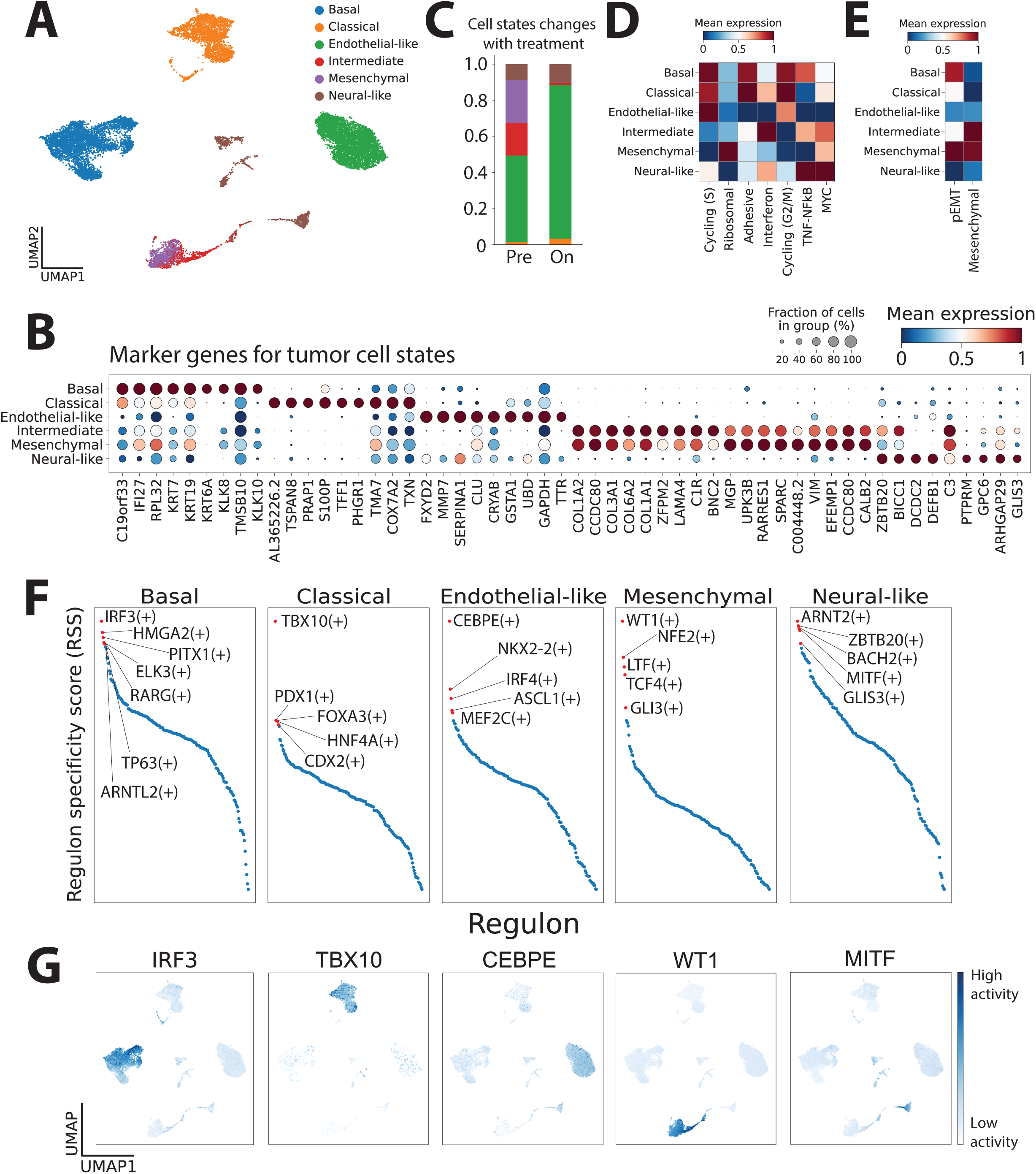
Defining novel tumor cell states in biliary tract cancers. (A) UMAP embedding of all tumor cells profiled by scRNAseq labeled by tumor cell states. (B) Dot plot showing marker genes for each tumor cell state. Differential expression analysis was performed using a Wilcoxon rank-sum test. (C) Proportion of each tumor cell state relative to all tumor cells in pre- and on-treatment biopsies. (D)-(E) Pathway scoring of (D) known cell cycle and signaling and (E) EMT gene programs in tumor cells belonging to each cell state. (F) Prediction of regulons (using SCENIC+^86^) with highest specificity in each cell state. (G) UMAP embedding of all tumor cells showing activity of select tumor cell state-specific regulons.

To better characterize the unidentified state (cNMF program 20), we performed gene-set enrichment analysis on the top 50 weighted genes in this program and found enrichment in pathways related to angiogenesis, blood vessel development, and morphogenesis (**Figure S10B**). We subsequently labeled this novel state ‘endothelial-like’ given the clear expression of genes related to blood vessel formation and endothelial cells (**Figure 3A,B**). Interestingly, we found the highest DKK1 expression and FZD gene program expression in classical cells (**Figure S10C**). CKAP4 was most highly expressed in mesenchymal cells, the Wnt gene signature was highest in basal cells, and the LRP gene signature was highest in neural-like cells (**Figure S10D,E**). To test the validity of our tumor cell states, we obtained an independent publicly available dataset^24^ of tumor cells obtained from intrahepatic cholangiocarcinoma and hepatocellular carcinoma biopsies. We scored tumor cells for gene programs for the five tumor states we identified in our dataset (**Figure 3A**) and observed expression of all five states in this validation dataset, albeit at different frequencies than observed in our data (**Figure S11**).

Across treatment timepoints, we interestingly noted a trend towards enrichment of endothelial-like state after treatment (**Figure 3C**), which may be suggestive of a role in treatment resistance. To better characterize these tumor cell states, we scored all tumor cells with several known tumor gene programs expressed, including cell cycle, adhesion, inflammatory signaling and EMT and partial EMT (**Figure 3D,E**). Of note, we found strong enrichment of adhesive programs in classical cells as might be expected. Interestingly we found that endothelial-like tumor cells had the strongest expression of S-phase cell cycling genes, suggestive of active cell cycle progression. Neural-like tumor cells had the strongest expression of TNF and MYC signaling, and intermediate cells had the strongest interferon signaling (**Figure 3D**). We additionally found basal cells expressed a recently described partial EMT program^60^, whereas mesenchymal cells had expression of both mesenchymal and partial EMT programs (**Figure 3E**).

### Identifying regulators of tumor cell states

To better understand what controls the expression of the five tumor cell programs we identified, we performed gene-regulatory network inference and regulon prediction^61^ on the scRNAseq data to identify transcriptional regulators of each tumor cell state. Interestingly, we found several validated and novel regulators within each state with differential expression profiles (**Figures 3F,G and S12**). We identified a TP63 regulon as one of the top regulons driving basal expression programs, consistent with its role in basal and squamous states across many tumor types^28,62^. Interestingly, the top two regulons driving basal expression programs were driven by a key interferon regulatory factor, IRF3, that drives type I interferon production, and by HMGA2, which is known to play a role in mesenchymal differentiation^63^. Among the regulons driving classical states were those driven by TBX10, PDX1, FOXA3 and CDX2, which is a known prognostic marker in gastrointestinal cancers, especially colorectal cancer, and has been described as a lineage survival oncogene^63,64^. Within mesenchymal cells, regulons driven by WT1, NFE2, TCF4 and GLI3 were among the top predictions. Consistent with these findings, a TCF4 regulatory network has recently been described to drive mesenchymal signatures and immunotherapy resistance in melanoma^65^.

Neural-like PDAC cells were recently described in a cohort of neoadjuvant treated PDAC patients and have been shown to be associated with chemoradiation resistance^28^. In our data, we confirmed a recent report that GLIS3 drives a neural-like malignant state in PDAC and proposed several novel regulators of this state including MITF, BACH2 and ARNT2 that may warrant further testing in BTCs. Within the novel endothelial-like state, we identified regulons driven by CEBPE, NKX2-2, IRF4, ASCL1, and MEF2C as important for program expression. Of note, ASCL1 has been described in several studies as a lineage defining factor for neuroendocrine cells in other tumors^66,67^ and also represses neural-crest like states via SOX9^68^, but it remains unclear what its connection may be in defining endothelial-like gene expression programs.

### Tumor cell states associate with distinct tumor immune cell types

Next, we sought to better understand how the TME may vary with cancer cells in each of the five identified cell states. To do this, we examined the co-variation of gene scores for each of the tumor programs within tumor cells with T cell and myeloid programs obtained by cNMF (**Figures S7 and S8**) within their respective cell types (**Figure 4**). We found that the mesenchymal tumor cell program covaried with the self-renewing immunosuppressive monocyte myeloid programs, and naive/memory, Gzmk and Tc17 T cell programs. This association may suggest a more tumor suppressive TME for mesenchymal cells, and may be consistent with known therapy resistance phenotypes for this cell state. We found tumor basal cell programs covaried with myeloid ISG and T cell ISG and proliferation programs. This therefore suggests that basal cells exist in an immune activated, interferon-rich TME. Interestingly, endothelial-like tumor programs covaried with myeloid proliferation and Treg programs, which may suggest these endothelial-like cells exist in a distinct suppressive TME. Together these data lay the groundwork for future studies to investigate potentially distinct TME niches for different cancer cell states.

**Figure 4.**
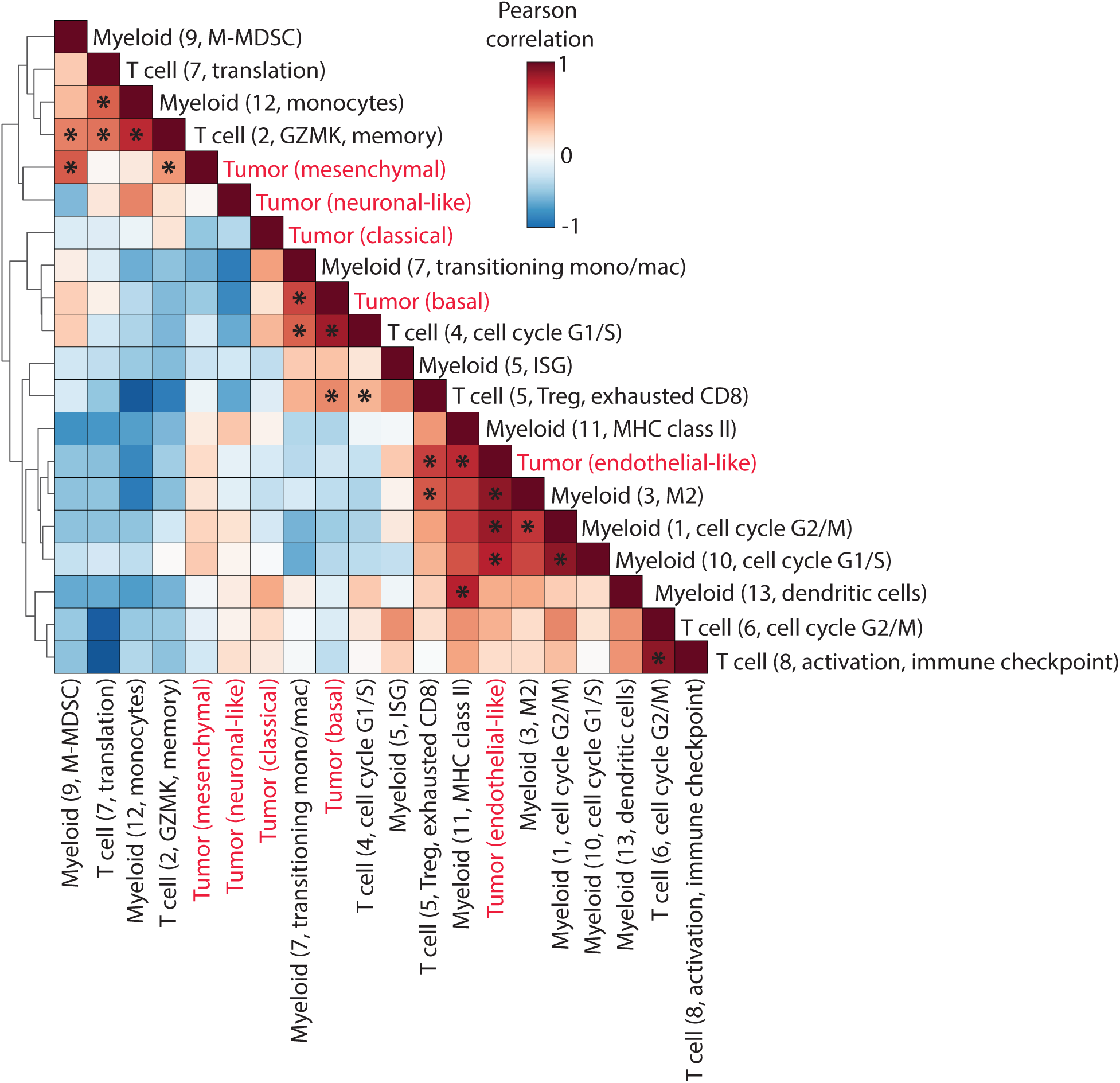
Heatmap showing Pearson correlation in abundance of all tumor cell states, T cell subsets and myeloid cell subsets across all samples.

## Discussion

We performed a phase II study combining DKN-01 with nivolumab in patients with biliary tract cancer (BTC) previously treated with systemic therapy. No objective responses were seen, highlighting the treatment refractory nature of this disease. Tumors exhibited low expression of DKK1 and PD-L1 across the cohort, which may have contributed to the absence of clinical activity.

In treatment-refractory tumors, molecular subtyping has revealed new modes of therapeutic resistance as well as subtype-specific vulnerabilities. Harnessing this knowledge, molecular subtype-guided treatment selection has shown promise in several aggressive cancers including pancreatic cancer^69^, treatment refractory triple-negative breast cancer^70^ and multiple other cancers included in the umbrella NCI-MATCH trial^71^. Most molecular subtyping, including in BTCs, has relied largely on bulk genomic and transcriptomic profiling^35,72^. While potentially targetable genetic alterations are found in ∼30% of BTCs^72,73^, targeting these alterations with IDH and FGFR inhibitors have shown efficacy only in a minority of patients^74–76^. A limitation of bulk molecular profiling is that it obscures intratumoral heterogeneity at the level of tumor cells and the TME, and misses rare cell populations that can mediate drug resistance. Single-cell RNA-seq addresses these limitations and can highlight novel mechanisms of treatment response and resistance as has been shown in pancreatic ductal adenocarcinoma (PDAC) and colorectal cancer^28,77–80^.

To explore mechanisms of treatment resistance in our trial and inform future clinical trials, we performed scRNAseq of pre-treatment and paired on-treatment tumor biopsies, and we deeply characterized malignant and immune cell states. We provide the first systematic classification of functionally annotated cell states in BTC based on characterization of five discrete malignant gene expression programs: classical, basal, mesenchymal, neural-like, and endothelial-like. Uniquely in our study, the first four of these malignant cell states have been identified and deeply characterized in PDAC, raising the potential of shared therapeutic resistance and vulnerabilities between these cancers with closely related ontogeny for their cell of origin.

The classical program has been extensively described in PDAC and is composed of epithelial differentiation genes. Pdx1, the master transcription factor for pancreatic epithelial development, was specifically active in classical-type cells (**Figures 3F and S12**), which exhibited high activity of a cell adhesion program (**Figure 3D**). The basal program is associated with invasion, metastasis, and treatment resistance across many solid tumors. p63 (encoded by TP63), a known regulator of basal cell identity, is specifically active within basal-type cells in BTC (**Figures 3F and S12**). Despite worse prognosis, basal-like breast^81^ and bladder^82^ cancers have enhanced chemosensitivity, supporting increased use of chemotherapy in these subtypes. Basal and mesenchymal subtypes have often been confounded in bulk studies. We found these to be distinct programs in BTC, consistent with a recent single-cell pan-cancer analysis^60^. Multiple regulators of epithelial-to-mesenchymal transition (EMT) were predicted as transcription factors (**Figure 3F**) for the mesenchymal but not basal program, including WT1, TCF4, and GLI3. The basal subtype demonstrated a partial EMT (pEMT) (e.g. vimentin, laminins) but not complete EMT (mesenchymal) signature (e.g. collagens)^60^ (**Figure 3E**) while the mesenchymal subtype had high expression of both.

We also discovered a neural-like malignant program in BTC, enriched in genes that regulate neuronal development and migration. The neural-like program was recently reported in PDAC^28^, is enriched in patient tumors treated with chemotherapy and radiation therapy, and may drive perineural invasion (PNI), which confers poor prognosis in PDAC and BTC^83^. GLIS3 was recently suggested to regulate the neural-like state in PDAC and is one of the top predicted transcription factors in our analysis of BTC (**Figure 3F**). Importantly, we identified a subset of malignant cells with high activity of both mesenchymal and neural-like programs, suggesting possible plasticity between these states. We found that an endothelial program is active in a subset of BTC malignant cells, which also uniquely express high levels of MMP7 (**Figure 3B**), a mediator of tumor invasion and metastasis, and exhibited high cell cycling activity (**Figure 3D**). These cells were also enriched in on-treatment samples (**Figure 3C**), suggesting resistance to the trial therapy. Endothelial transdifferentiation of malignant cells was first described in melanoma^84^ and, consistent with our findings, facilitates tumor progression under hypoxia and resistance to anti-angiogenic therapies. Our analysis predicted CEB/P epsilon, an essential regulator of granulocyte differentiation, as a selective regulator of this program, which will need to be validated in follow-up studies.

Emerging evidence suggests that cancer cell states colocalize with differing immune and stromal cell types to form distinct neighborhoods^28,60,85^, and these multicellular interactions mediate treatment responsiveness. We find distinct multicellular hubs that are characterized by varying forms of immune suppression, such as mesenchymal cancer cells associated with self-renewing, immunosuppressive monocyte and GzmK/progenitor T cells and endothelial-like cancer cells associated with “M2” immune suppressive myeloid programs as well as regulatory and exhausted T cells (**Figure 4**). Expansion to a larger cohort and spatial data will be necessary to validate these multicellular hubs and determine their prognostic and predictive significance to guide precision therapies.

Analysis of on-treatment vs pre-treatment biopsies identified changes in all cellular compartments induced by trial therapy. Enrichment of endothelial-like tumor cells after immunotherapy (**Figure 3C**) may be due to the co-localization of M2-polarized macrophages and regulatory T cells that we observed. Immune cell dynamics reflected increased immune activation including expansion of exhausted CD8 T cells, ISG+ CD4 T cells, resident memory T cells, and a shift from monocytes to macrophages. However, immune suppressive gene expression changes occurred within the immune cells, including M2 signature upregulation within proliferating myeloid cells, downregulated antigen presentation in both dendritic cells and macrophages, and upregulated chronic exhaustion signatures in resident memory CD4 and CD8 T cells and exhausted CD8 T cells.

In summary, we provide here an initial framework for the high-resolution classification of biliary tract cancers and characterization of their response to therapy. Through analysis of gene expression modules in malignant and immune cells, we made preliminary insights into the biology that drives aggressiveness and treatment resistance to immune checkpoint inhibitors in BTC. The neural-like program, which may be a key driver for perineural invasion, has been functionally validated in other cancers but reported for the first time here in BTC. Endothelial transdifferentiation of malignant cells is functionally implicated in resistance to antiangiogenic therapy in other cancers and described in our study for the first time in BTC. We characterized the immune TME associated with each malignant cell state and identified multiple pathways of immune evasion and therapeutic resistance. While a limitation of our study is the small number of paired samples, our analysis of this curated sample set provides a framework for analyzing treatment resistance mechanisms and therapeutic vulnerabilities in individual tumors. Spatial analyses and larger prospective cohorts will be necessary to validate the TME subtypes and treatment-associated changes and potential therapeutic resistance mechanisms we have identified. This will yield prognostic and predictive information about each subtype that can be harnessed for patient stratification, biomarker selection, and therapeutic selection for the next generation of trials in BTC.

### Patients and Methods

#### Study design

This single-arm, multicenter, phase II study was designed to assess the efficacy of the combination of DKN-01 and nivolumab in previously treated patients with advanced biliary tract cancers (BTCs). The primary endpoint was objective response rate (ORR). The secondary endpoints included 1) progression free survival (PFS), 2) median overall survival (OS), 3) ORR, PFS and OS of subgroups stratified by tumor DKK1 and PD-L1 expression level, 4) tolerability and safety and 5) assessment of the biological effects of target inhibition in correlative studies. Patients were enrolled under DFCI/MGH IRB protocol #19-200 and treated between October 2019 and August 2020. Baseline biopsies were performed for each patient prior to the first treatment on trial by the interventional radiology (IR) team at MGH. On-treatment biopsies were performed at 3-weeks into the trial protocol by the IR team. Core biopsies obtained during this study were subsequently processed as described below.

#### Immunohistochemistry

For the evaluation of PD-L1 protein expression, FFPE sections were deparaffinized by baking them for 1 hour at 60°C. IHC staining was done on the BondRx using the BOND Polymer Refine Detection kit (Catalogue No. DS9800). Antigen retrieval was carried out at pH 7.5 for 20 mins using Bond Epitope Retrieval Solution 2 (Leica Biosystems). PD-L1 Rabbit monoclonal E1L3N antibody was purchased from Cell Signaling Technology (Catalogue No. 13684S) and used at a concentration of 1:200.

#### RNA in chromogenic situ hybridization

Automated DKK1 RNAscope chromogenic in situ hybridization (CISH) was performed using the 2.5 LS Reagent Kit-Red from Advanced Cell Diagnostics (ACD) (Catalogue No.322150) on the BondRx platform. 5-μm sections of FFPE tissue were mounted on Surgipath X-tra glass slides, baked for 1 hour at 60°C, and placed on the BondRx for processing. On the BondRx, the staining protocol used was the ACD ISH RED Protocol. The RNA unmasking conditions for the tissue consisted of a 15-minute incubation at 95°C in Bond Epitope Retrieval Solution 2 (Leica Biosystems) followed by 15-minute incubation with Proteinase K which was provided in the kit. Probe hybridization was done for 2 hours with RNAscope probes which were provided by ACD. The probes used for this study were, LS 2.5 Hs-DKK1 Probe (Catalogue No. 421418); and LS 2.5 Positive Control Probe_Hs-PPIB (Catalogue No. 313908) which was used as a housekeeping control probe to assess RNA integrity.

#### Generation of single-cell RNA-sequencing data

Fresh tissue biopsy cores were dissociated using the human tumor dissociation kit (Miltenyi Biotec; 130-095-929). Tissue was first minced into small pieces using a scalpel, and placed in a 1.5mL eppendorf tube containing enzyme H (100uL), enzyme R (50uL), enzyme A (12.5uL) and RPMI (837.5uL). The sample was incubated for 20 minutes in a thermomixer at 37degC at 600rpm. The sample was subsequently strained with a 70uM cell strainer and washed with cold PBS containing 1.5% FBS. Samples were subsequently spun down at 1300rpm in 4degC for 5 minutes, and resuspended in PBS for cell counts and to assess viability with trypan blue using a Countess automated cell counter (Invitrogen). Samples were subsequently processed using the 10X 5’ V1 protocol as per manufacturer’s instructions. Briefly, for each sample ∼10,000 cells were loaded onto a 10X chromium controller for GEM generation to recover ∼6,000 cells. Samples were subsequently processed for GEM breakage, cDNA amplification and library preparation as outlined in the protocol. Final libraries were quantified using a bioAnalyzer and pooled for sequencing on a NovaSeq S4 machine at a sequencing depth of ∼50,000 reads per cell.

#### Processing of single-cell RNA-sequencing data

Raw sequencing files were demultiplexed using Cellranger (v6.1.2) mkfastq and count matrices were generated using Cellranger (v6.1.2) counts with alignment to the GRCh38-2020-A reference genome. All analysis of the scRNAseq data was performed in Python (version 3.7.10). The filtered count output from the cellranger count pipeline per sample was concatenated using the Python package Scanpy (https://github.com/theislab/scanpy). The count matrix was further filtered to remove cells with less than 50 genes expressed, less than 200 total counts, or over 25% mitochondrial gene counts, and to remove genes that are expressed in at least 3 cells. The Python package DoubletDetection (https://github.com/JonathanShor/DoubletDetection) was run to infer and remove doublets on a per sample basis. The count matrix was then normalized to 1000 total counts per cell (scanpy.pp.normalize_total) and logarithmized (scanpy.pp.log1p). The highly variable genes are calculated (scanpy.pp.highly_variable_genes, mean range of 0.0125 to 3, minimum dispersion of 0.5). Principal component analysis is then performed (scanpy.tl.pca), and the top 40 components are used to generate a neighborhood graph (scanpy.pp.neighbors). A UMAP was generated and leiden clustering was performed at a resolution of 0.5 (scanpy.tl.umap, scanpy.tl.leiden). A set of known marker genes^36^ was used to identify and annotate the cell types of each leiden cluster. The top ranked genes per leiden cluster were also identified using a t-test, wilcoxon rank sum test, and logistic regression (scanpy.tl.rank_genes_groups), and these gene lists were used to further confirm the annotations.

#### Copy-number inference

InferCNV (https://github.com/broadinstitute/infercnv) was run on the unprocessed epithelial cells from all samples to infer copy number variations. The inputs include the combined count matrix, cell type annotations per cell barcode, and the list of cell types to be used as reference. A gene ordering file was generated from the standard hg38 genome reference using the script provided in the above github repo. The i6 HMM implementation of inferCNV was used with a cutoff of 0.1 (as advised for 10x data), the subclusters analysis mode and median filtering enabled. A score was calculated from the HMM state for each inferCNV cluster by centering to zero and taking the sum of the squared value for each region. This cluster level score was then applied to each cell belonging to the cluster and was used to identify malignant cells.

#### Consensus non-negative matrix factorization

cNMF (https://github.com/dylkot/cNMF) was run on each cell type separately to derive gene programs. For each of these runs, the input was an unprocessed AnnData file subset to that cell type only. The custom parameters used include the number of highly variable genes to use in the factorization step (2000) and the range of k values tested for by cNMF (5 to 101 with intervals of 3). All other default parameters were used. After the prepare, factorization, and combine steps were completed, the k selection plot was used to manually select the most appropriate k value, or number of factors, and then run the consensus step.

#### Inference of regulatory networks

pySCENIC (https://pyscenic.readthedocs.io/en/latest/index.html) was run on the tumor cells to infer transcription factors and regulons. The AnnData file was subset to just contain tumor cells and then converted into the required loom file. The GRNBoost2 algorithm was used to infer gene regulatory networks given the loom file and default transcription factor list. This outputs a matrix containing a list of adjacencies between a transcription factor and its targets. Regulon inference (cisTarget) was performed using the default ranking and motif annotation databases, along with the same loom file and adjacencies matrix generated in the previous step. Cellular enrichment (AUCell) was calculated using the same loom file, the regulon matrix generated in the previous step, and all other default parameters. Downstream analysis was performed using the previously identified tumor cell subtype annotations. The regulons with the highest specificity scores for each tumor cell subtype were selected for further analysis.

#### Analysis of co-varying programs

To find sets of co-varying gene programs, a sample by program matrix was generated where the values were the 90th percentile of the cNMF program score for that program and sample. This was repeated for the 25th, 50th, 75th, and 95th percentiles as well. For each of these matrices a pairwise correlation of columns was calculated using pandas’ DataFrame.corr() method, resulting in a program by program correlation matrix. This was then visualized using Seaborn’s clustermap function with Euclidean distance as the spatial distance metric. To calculate significance, the sample assignments for each cell were randomly shuffled using pandas’ Series.sample() method and then the 75th percentile scores and correlation coefficients were recalculated. This was repeated for a 1000 iterations to generate a null distribution of correlations for each pair of programs. A p-value was then calculated by counting how many times the randomly shuffled patient assignment correlations were higher than the true correlation, divided by the total number of iterations.

#### Statistical analysis

All statistical analyses were performed using a student t-test with Bonferroni correction unless otherwise specified in the text.

## Data Availability

All data produced in the present study are available upon reasonable request to the authors

All single-cell RNA-sequencing data will be made publicly available at the time of publication. All code used for analysis will be made available at: https://github.com/arnav-mehta/cholangio-dkn01-nivo.

## Acknowledgements

We would like to thank funding support from the Prostate Cancer Foundation Young Investigator Award (RJP), NIH R01CA260872 (DGD), NIH R01CA260857 (DGD), NIH R01CA247441 (DGD), and DoD PRCRP / W81XWH-21-1-0738 (DGD), Dana Farber Cancer Institute / Harvard CancerCare GI SPORE Career Enhancement Award (AM), Sky Foundation Pancreatic Cancer Research Grant (AM), Doris Duke Charitable Foundation Physician Scientist Fellowship (AM), DF/HCC K12 (K12CA087723) Paul Calabresi Award for Clinical Oncology (AM), a American Cancer Society Clinical Scientist Development Grant (134013-CSDG-19-163-01-TBG) (LG) and the Cholangiocarcinoma Foundation Andrea Marie Fuquay Research Fellowship (LG). We additionally would like to thank for funding support from Leap Therapeutics who supported the experimental work performed in this study.

## Supplemental Figure Legends

**Figure S1.** InferCNV profile of all tumor cells against normal cell references within the scSeq dataset.

**Figure S2.** UMAP embedding of scRNAseq profiles of all cells obtained across biopsy samples labeled by (A) patient sample and (B) diagnosis histology. (C) Cell type proportions of all major cell types obtained by scRNAseq split by sample.

**Figure S3.** (A) Table showing number and percentage of cells of each major cell subset that were DKK+. Scoring of (B) DKK1 ISH and (C) PD-L1 IHC staining on a subset of samples for which FFPE tissue was available. (D) Violin plots showing expression of DKK1, LRP6, a FZD gene signature and WNT gene signature split but major cell types. (E) Proportion of cells belonging to each major cell type split by cells that were DKK1+ and DKK1-. (F) Dot plot showing Wnt signature scoring across all major cell types split by pre- and on-treamtent.

**Figure S4.** (A) UMAP embedding of all NK cells obtained by scRNAseq labeled by granular subsets. (B) Heatmap showing marker genes for NK cell subsets.

**Figure S5.** Dot plot showing CKAP4 expression in all (A) major cell types, (B) CD4+ T cell subsets, (C) CD8+ T cell subsets, (D) myeloid subsets, and (E) NK cell subsets.

**Figure S6.** Dot plot showing WNT signature scoring stratified by pre- and on-treatment samples in (A) CD4+ T cell subsets, (B) CD8+ T cell subsets, (C) myeloid subsets, and (D) NK cell subsets.

**Figure S7.** (A) Gene programs identified by consensus non-negative matrix factorization (cNMF) on all T cells. Shown is a dot plot of the usage of each identified gene program within each myeloid subset. (B) Correlation heatmap of cNMF usages with known, annotated gene programs from an independent dataset (Pelka et al). (C) Changes over treatment timepoints in Usage 2 (GzmK), Usage 4 (proliferation), Usage 6 (proliferation) and Usage 8 (CXCL13) in CD4+ and CD8+ T cell subsets.

**Figure S8.** (A)-(B) Gene programs identified by cNMF on all myeloid cells. Shown is a dot plot of the usage of each identified gene program within each myeloid subset. (B) Correlation heatmap of cNMF usages with known, annotated gene programs from an independent dataset (Pelka et al). (C) Changes over treatment timepoints in Usage 5 (interferon stimulated genes), Usage 1 (proliferation) and Usage 10 (proliferation) in myeloid cell subsets.

**Figure S9.** (A) UMAP embedding of all tumor cells labeled treatment timepoint, biopsy tissue type, patient sample, diagnosis histology, and SPP1 or S100P expression level. (B) Gene programs identified by cNMF performed on all tumor cells. Shown is a dot plot of the usage of each identified gene program within tumor cells of each sample.

**Figure S10.** (A) UMAP embedding of all tumor cells showing expression of classical, basal, neural, mesenchymal and endothelial-like gene signatures. (B) Pathways enriched by gene-set enrichment analysis (GSEA) for top genes in the endothelial-like gene signature discovered by cNMF. Expression of (C) DKK1, (D) CKAP4, and (E) WNT, FZD and LRP gene signatures split by tumor cell states.

**Figure S11.** UMAP embeddings of an independent scSeq dataset of iCCA and HCC samples colored by Moffitt basal and classical signatures, and Hwang et al classical, neuronal-like and mesenchymal signatures, and Usage 20, which is our endothelial-like signature.

**Figure S12.** UMAP embeddings showing activity level of top predicted regulons in each tumor cell state.

## Supplemental Table Legends

**Table S1**. Table with baseline characteristics on all patients enrolled in the clinical trial.

**Table S2**. Table showing adverse events in the clinical trial.

**Table S3**: Table shows individual patient data including demographics, tumor characteristics, and response data

**Table S4**. Gene programs obtained by cNMF for myeloid and T cells.

**Table S5**. Gene programs for each tumor cell state obtained by cNMF in this study or from previous literature reports.

