## Supplemental Figures for "Characterization of cell states in biliary tract cancers identifies mechanisms of therapeutic resistance in a phase II trial of DKN-01/nivolumab"

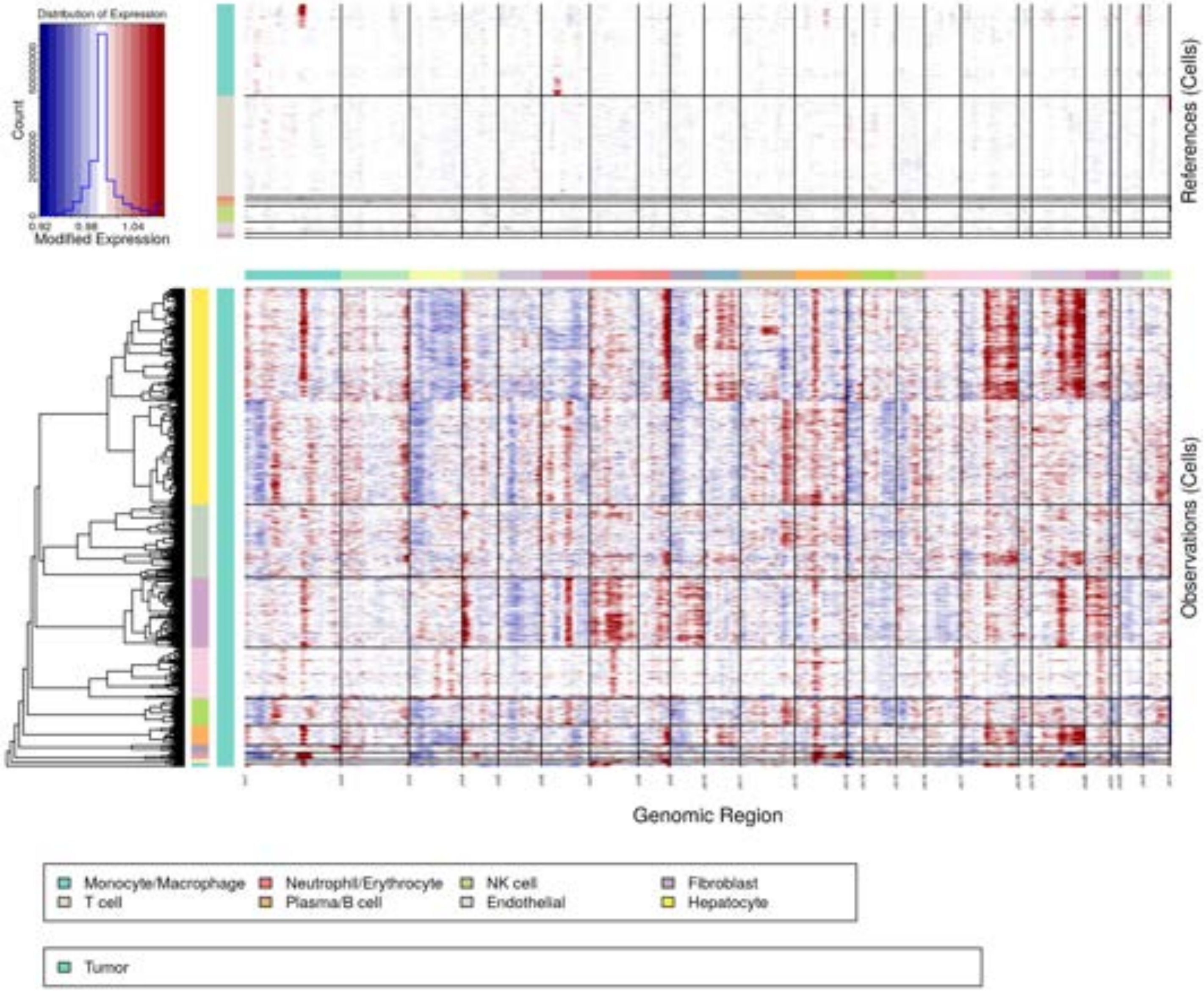
Distribution of Expression
Count
Modified Expression
References (Cells)
Observations (Cells)
Genomic Region
Monocyte/Macrophage
T cell
Neutrophil/Erythrocyte
Plasma/B cell
NK cell
Endothelial
Fibroblast
Hepatocyte
Tumor

Figure S1

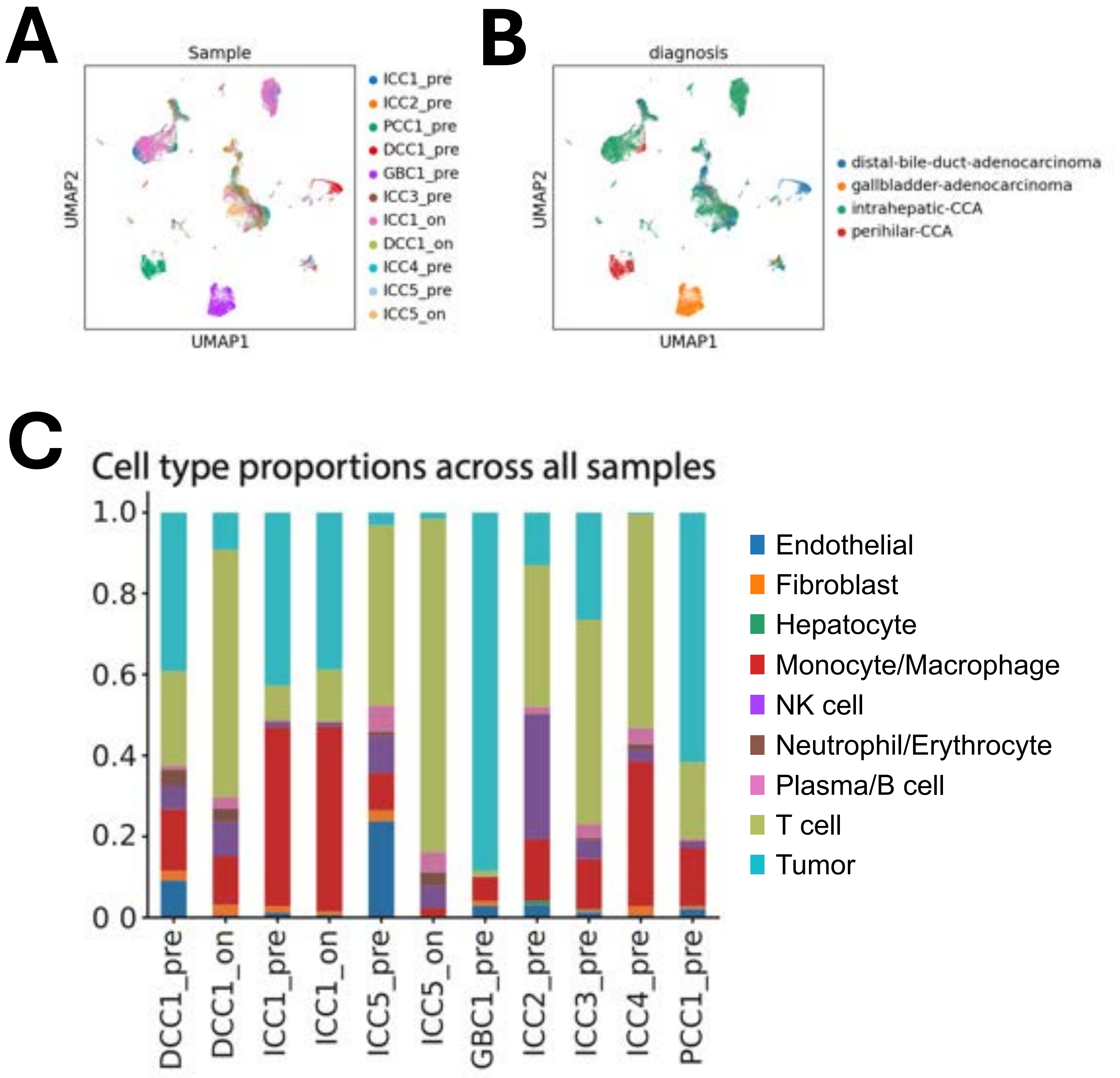
A
Sample
ICC1_pre
ICC2_pre
PCC1_pre
DCC1_pre
GBC1_pre
ICC3_pre
ICC1_on
DCC1_on
ICC4_pre
ICC5_pre
ICC5_on
UMAP2
UMAP1
B
diagnosis
distal-bile-duct-adenocarcinoma
gallbladder-adenocarcinoma
intrahepatic-CCA
perihilar-CCA
UMAP2
UMAP1
C
Cell type proportions across all samples
1.0
0.8
0.6
0.4
0.2
0.0
DCC1_pre
DCC1_on
ICC1_pre
ICC1_on
ICC5_pre
ICC5_on
GBC1_pre
ICC2_pre
ICC3_pre
ICC4_pre
PCC1_pre
Endothelial
Fibroblast
Hepatocyte
Monocyte/Macrophage
NK cell
Neutrophil/Erythrocyte
Plasma/B cell
T cell
Tumor

Figure S2

**A**

| Cluster | DKK1- | DKK1+ | % positive |
|---|---|---|---|
| Tumor | 18,299 | 811 | 4% |
| T cell | 12,952 | 9 | < 0.1% |
| Mono/Mac | 11,827 | 50 | 0.4% |
| NK cell | 2,056 | 1 | < 0.1% |
| Endothelial | 1,460 | 1 | < 0.1% |
| Plasma/B | 729 | 0 | < 0.1% |
| Fibroblast | 616 | 10 | 1.6% |
| Neutrophil/erythrocyte | 514 | 1 | 0.2% |
| Hepatocyte | 44 | 0 | < 0.1% |

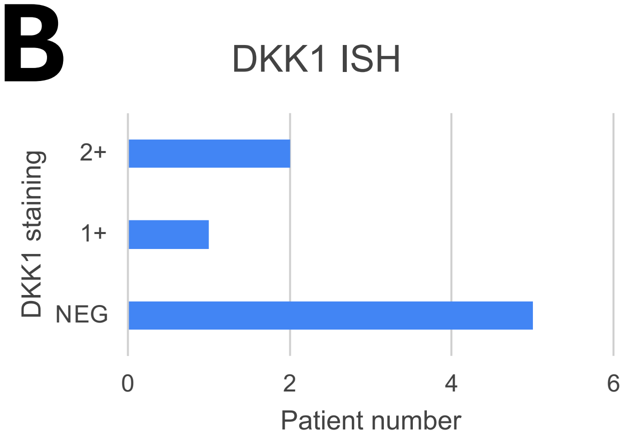


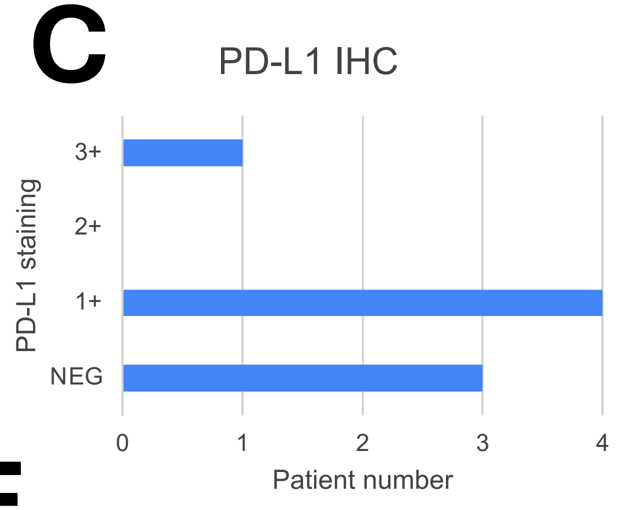


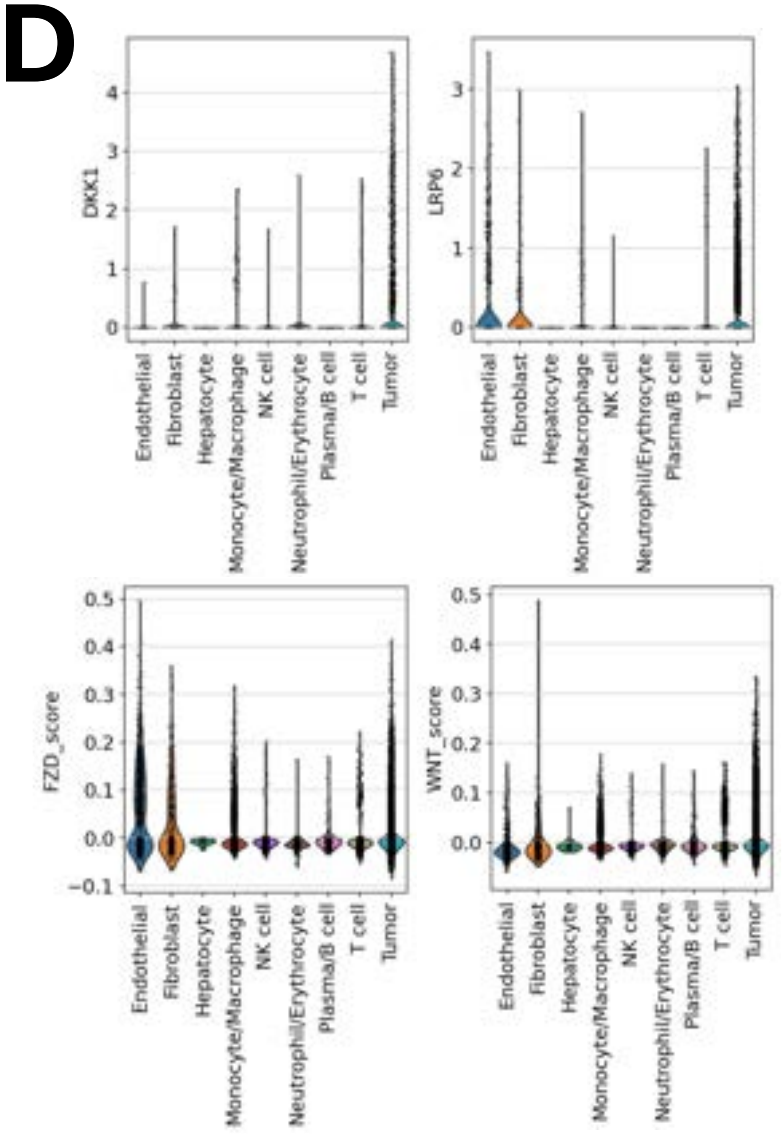


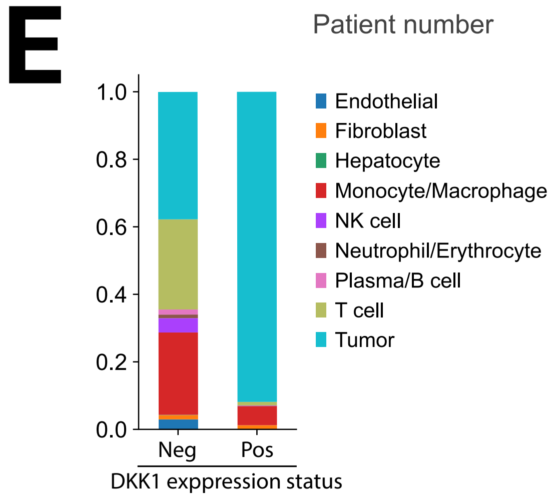


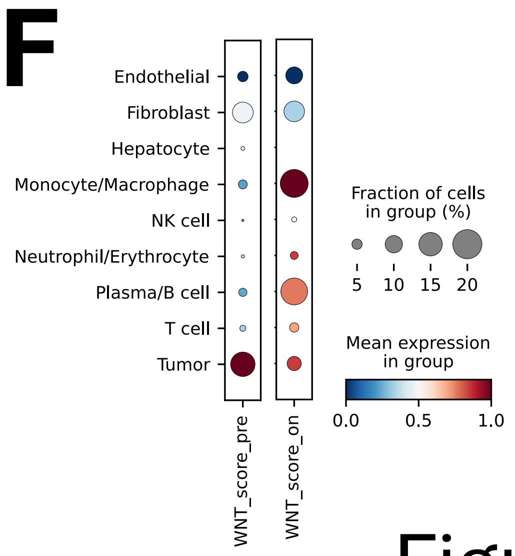


Figure S3

A

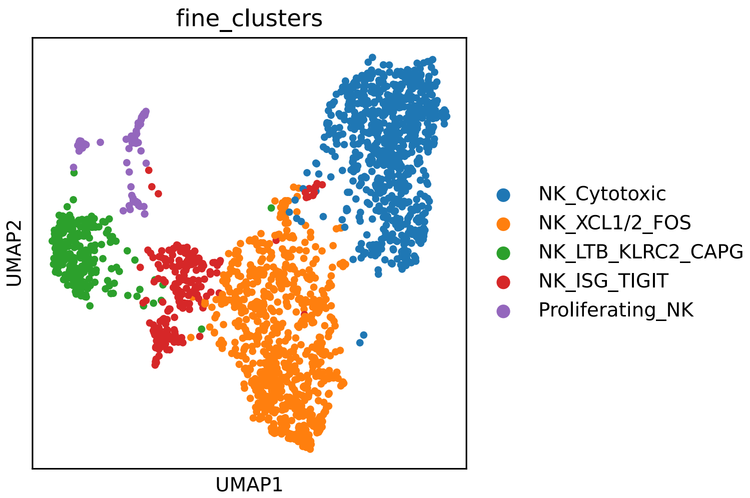

fine_clusters
UMAP2
UMAP1
NK_Cytotoxic
NK_XCL1/2_FOS
NK_LTB_KLRC2_CAPG
NK_ISG_TIGIT
Proliferating_NK


B

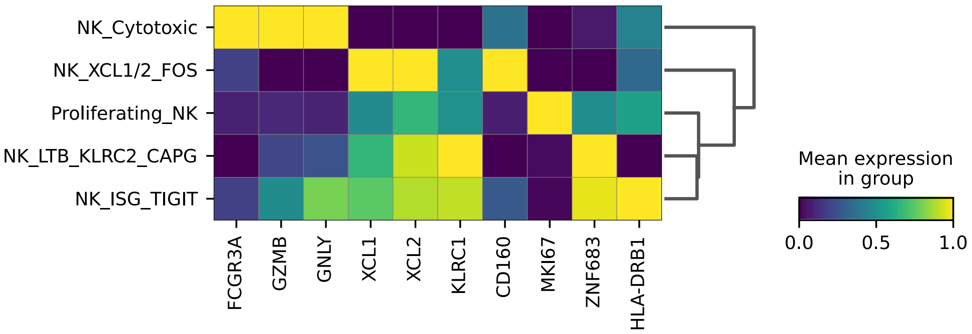

NK_Cytotoxic
NK_XCL1/2_FOS
Proliferating_NK
NK_LTB_KLRC2_CAPG
NK_ISG_TIGIT
FCGR3A
GZMB
GNLY
XCL1
XCL2
KLRC1
CD160
MKI67
ZNF683
HLA-DRB1
Mean expression in group
0.0
0.5
1.0


Figure S4

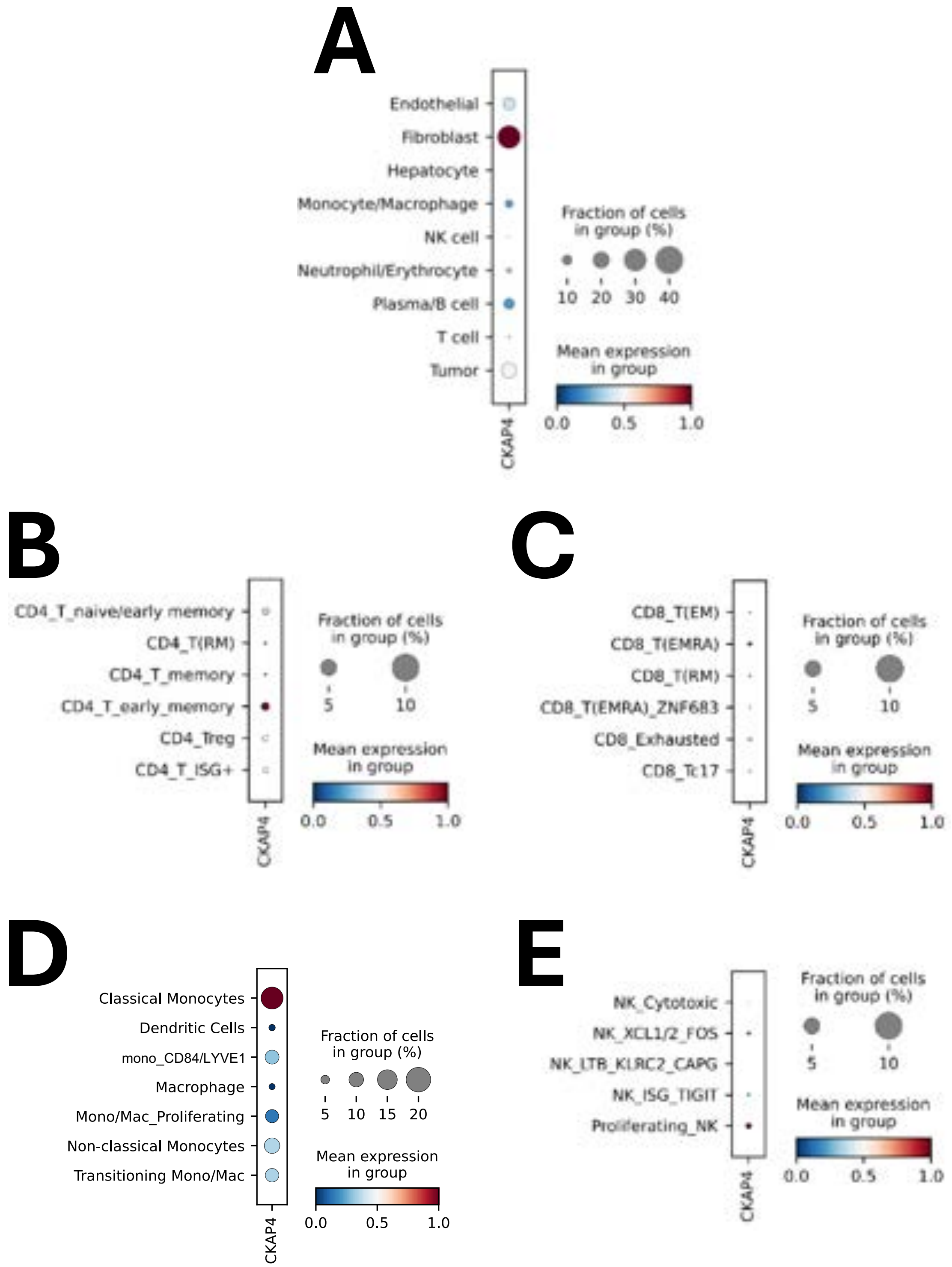
A
Endothelial
Fibroblast
Hepatocyte
Monocyte/Macrophage
NK cell
Neutrophil/Erythrocyte
Plasma/B cell
T cell
Tumor
CKAP4
Fraction of cells in group (%)
10 20 30 40
Mean expression in group
0.0 0.5 1.0
B
CD4_T_naive/early memory
CD4_T(RM)
CD4_T_memory
CD4_T_early_memory
CD4_Treg
CD4_T_ISG+
CKAP4
Fraction of cells in group (%)
5 10
Mean expression in group
0.0 0.5 1.0
C
CD8_T(EM)
CD8_T(EMRA)
CD8_T(RM)
CD8_T(EMRA)_ZNF683
CD8_Exhausted
CD8_Tc17
CKAP4
Fraction of cells in group (%)
5 10
Mean expression in group
0.0 0.5 1.0
D
Classical Monocytes
Dendritic Cells
mono_CD84/LYVE1
Macrophage
Mono/Mac_Proliferating
Non-classical Monocytes
Transitioning Mono/Mac
CKAP4
Fraction of cells in group (%)
5 10 15 20
Mean expression in group
0.0 0.5 1.0
E
NK_Cytotoxic
NK_XCL1/2_FOS
NK_LTB_KLRC2_CAPG
NK_ISG_TIGIT
Proliferating_NK
CKAP4
Fraction of cells in group (%)
5 10
Mean expression in group
0.0 0.5 1.0

Figure S5

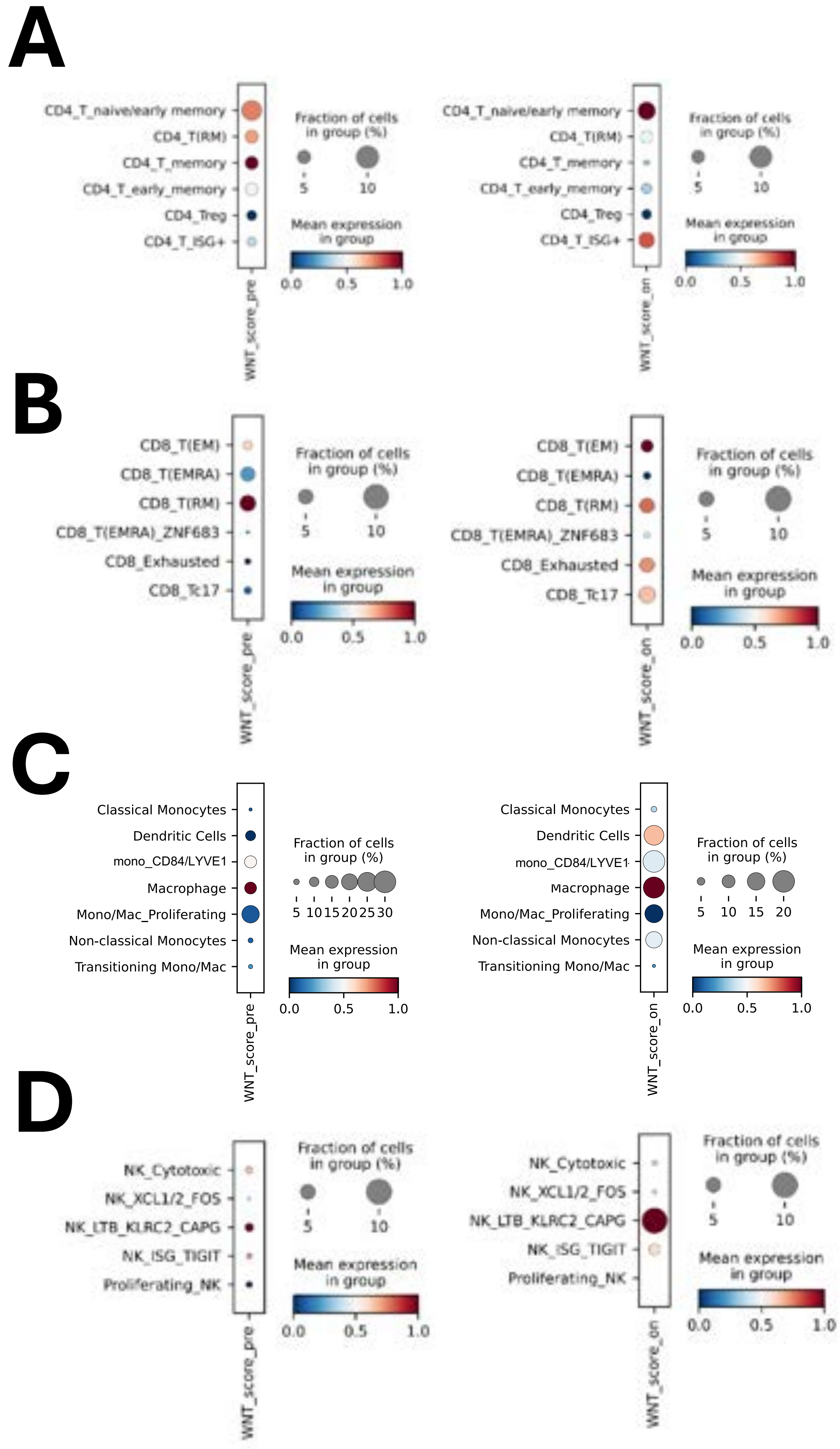
A
CD4_T_naive/early memory
CD4_T(RM)
CD4_T_memory
CD4_T_early_memory
CD4_Treg
CD4_T_ISG+
WNT_score_pre
WNT_score_on
Fraction of cells in group (%)
5
10
Mean expression in group
0.0
0.5
1.0
B
CD8_T(EM)
CD8_T(EMRA)
CD8_T(RM)
CD8_T(EMRA)_ZNF683
CD8_Exhausted
CD8_Tc17
C
Classical Monocytes
Dendritic Cells
mono_CD84/LYVE1
Macrophage
Mono/Mac_Proliferating
Non-classical Monocytes
Transitioning Mono/Mac
5 10 15 20 25 30
5 10 15 20
D
NK_Cytotoxic
NK_XCL1/2_FOS
NK_LTB_KLRC2_CAPG
NK_ISG_TIGIT
Proliferating_NK

Figure S6

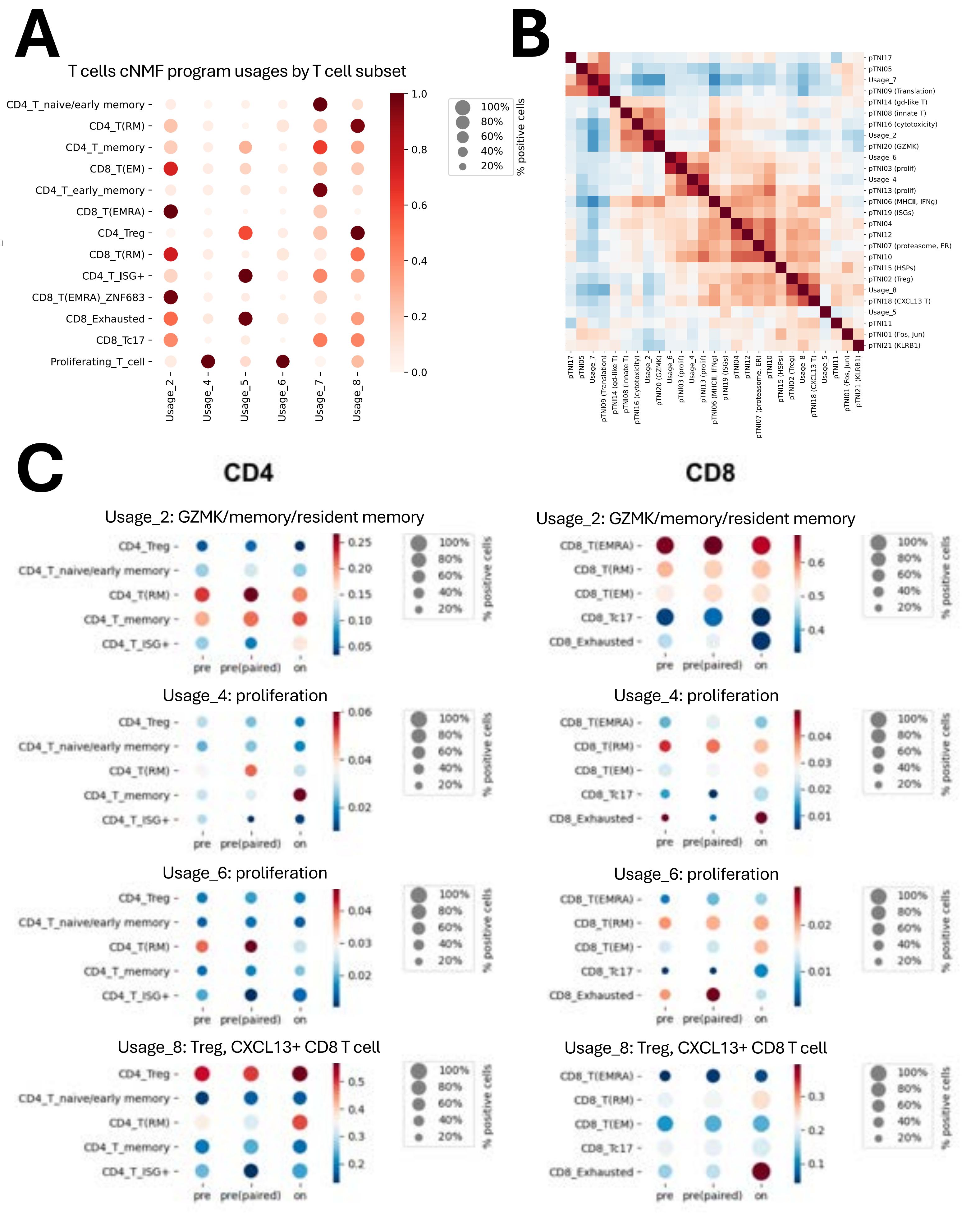
A
T cells cNMF program usages by T cell subset
CD4_T_naive/early memory
CD4_T(RM)
CD4_T_memory
CD8_T(EM)
CD4_T_early_memory
CD8_T(EMRA)
CD4_Treg
CD8_T(RM)
CD4_T_ISG+
CD8_T(EMRA)_ZNF683
CD8_Exhausted
CD8_Tc17
Proliferating_T_cell
Usage_2
Usage_4
Usage_5
Usage_6
Usage_7
Usage_8
% positive cells
B
C
CD4
CD8
Usage_2: GZMK/memory/resident memory
Usage_4: proliferation
Usage_6: proliferation
Usage_8: Treg, CXCL13+ CD8 T cell
pre
pre(paired)
on

Figure S7

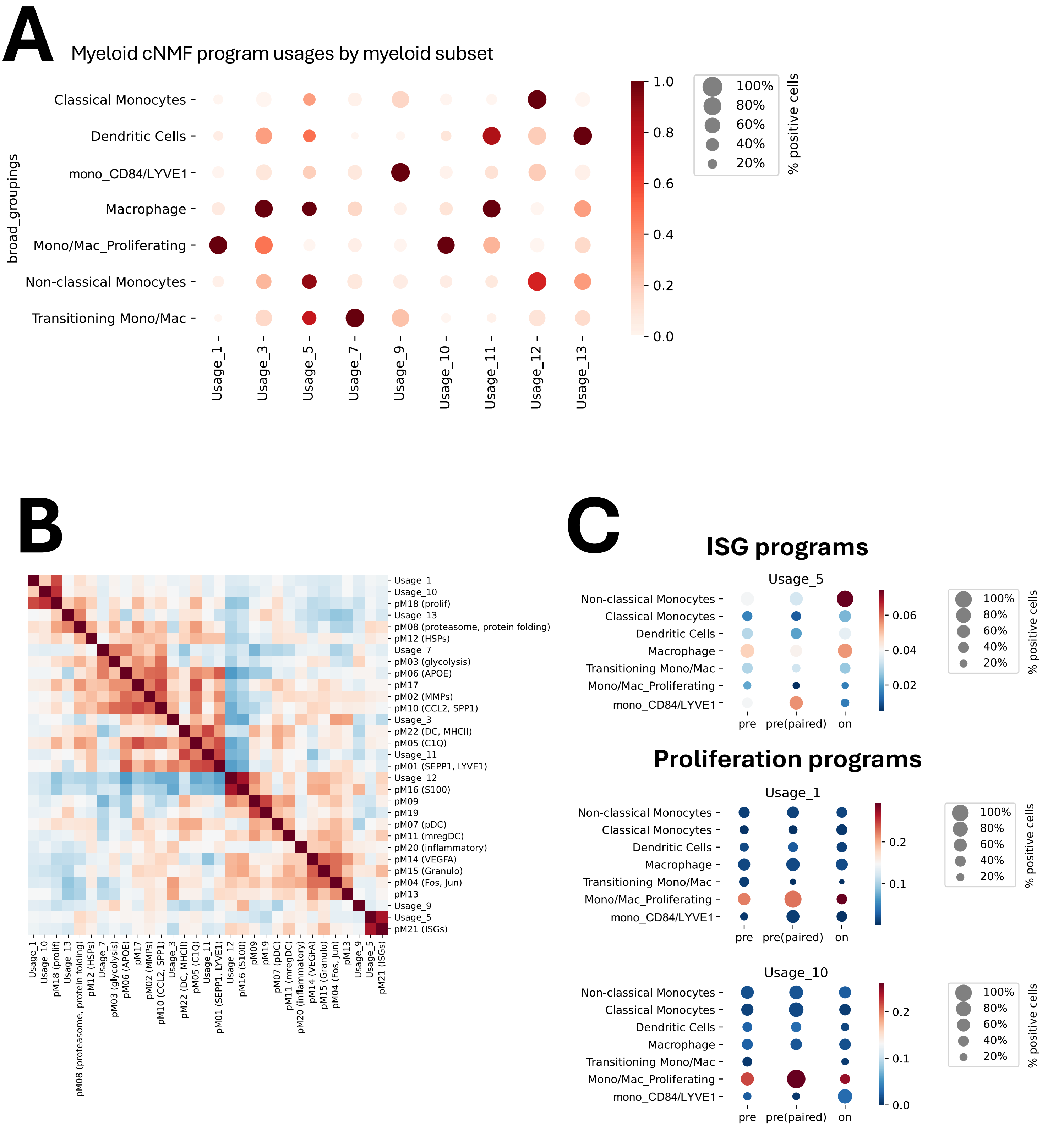

A
Myeloid cNMF program usages by myeloid subset
broad_groupings
Classical Monocytes
Dendritic Cells
mono_CD84/LYVE1
Macrophage
Mono/Mac_Proliferating
Non-classical Monocytes
Transitioning Mono/Mac
Usage_1
Usage_3
Usage_5
Usage_7
Usage_9
Usage_10
Usage_11
Usage_12
Usage_13
1.0
0.8
0.6
0.4
0.2
0.0
100%
80%
60%
40%
20%
% positive cells
B
Usage_1
Usage_10
pM18 (prolif)
Usage_13
pM08 (proteasome, protein folding)
pM12 (HSPs)
Usage_7
pM03 (glycolysis)
pM06 (APOE)
pM17
pM02 (MMPs)
pM10 (CCL2, SPP1)
Usage_3
pM22 (DC, MHCII)
pM05 (C1Q)
Usage_11
pM01 (SEPP1, LYVE1)
Usage_12
pM16 (S100)
pM09
pM19
pM07 (pDC)
pM11 (mregDC)
pM20 (inflammatory)
pM14 (VEGFA)
pM15 (Granulo)
pM04 (Fos, Jun)
pM13
Usage_9
Usage_5
pM21 (ISGs)
C
ISG programs
Usage_5
Non-classical Monocytes
Classical Monocytes
Dendritic Cells
Macrophage
Transitioning Mono/Mac
Mono/Mac_Proliferating
mono_CD84/LYVE1
pre
pre(paired)
on
0.06
0.04
0.02
100%
80%
60%
40%
20%
% positive cells
Proliferation programs
Usage_1
0.2
0.1
Usage_10
0.2
0.1
0.0


Figure S8

A

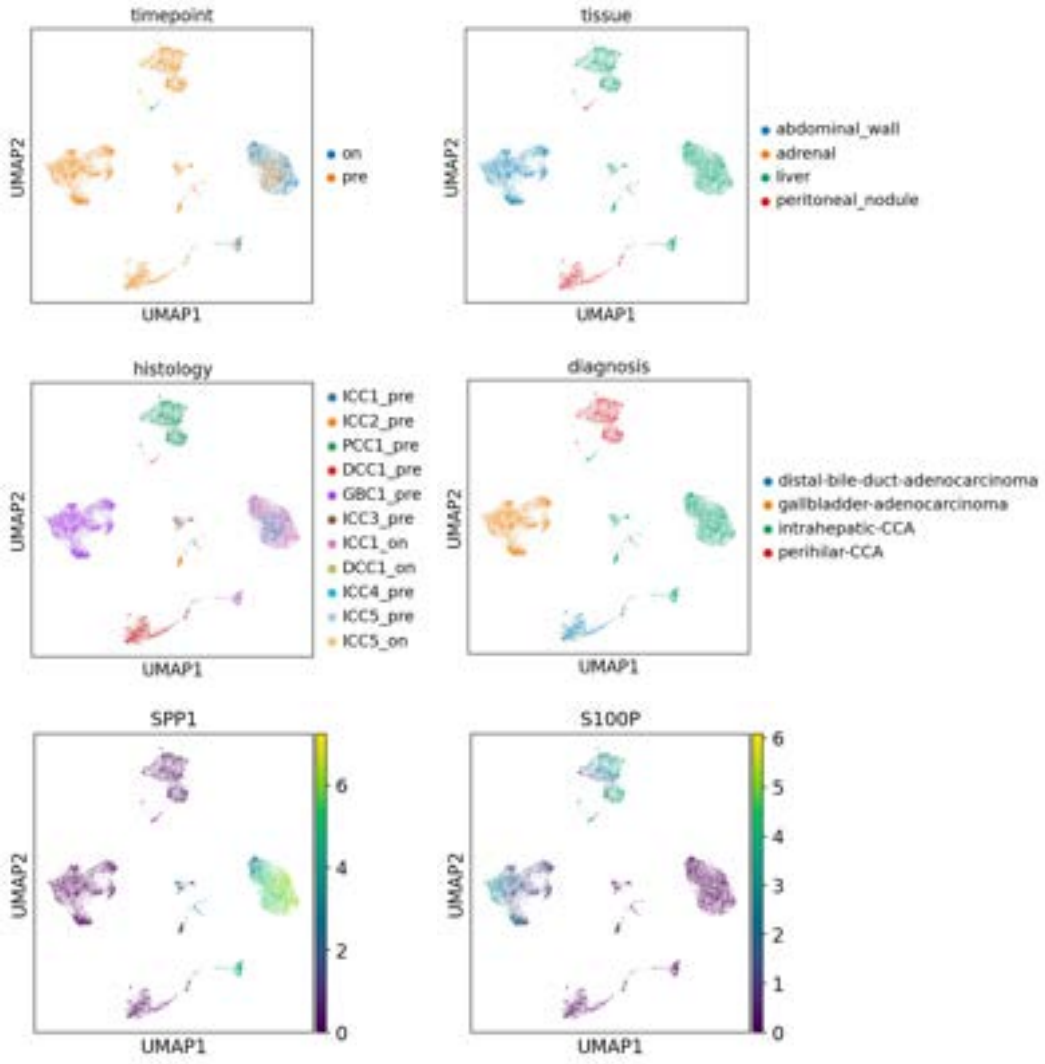
timepoint
tissue
histology
diagnosis
SPP1
S100P
UMAP1
UMAP2
on
pre
abdominal_wall
adrenal
liver
peritoneal_nodule
ICC1_pre
ICC2_pre
PCC1_pre
DCC1_pre
GBC1_pre
ICC3_pre
ICC1_on
DCC1_on
ICC4_pre
ICC5_pre
ICC5_on
distal-bile-duct-adenocarcinoma
gallbladder-adenocarcinoma
intrahepatic-CCA
perihilar-CCA

B

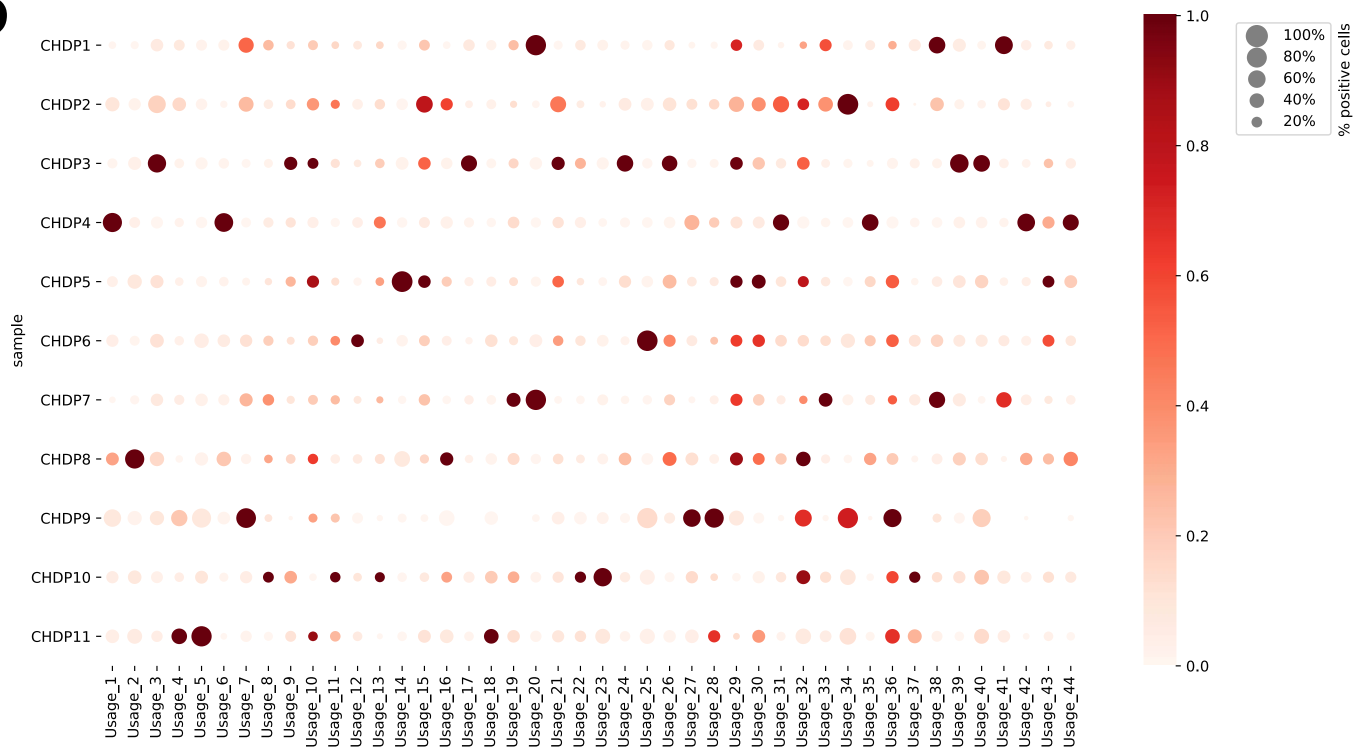
CHDP1
CHDP2
CHDP3
CHDP4
CHDP5
CHDP6
CHDP7
CHDP8
CHDP9
CHDP10
CHDP11
sample
Usage_1 … Usage_44
100%
80%
60%
40%
20%
% positive cells
1.0
0.8
0.6
0.4
0.2
0.0

Figure S9

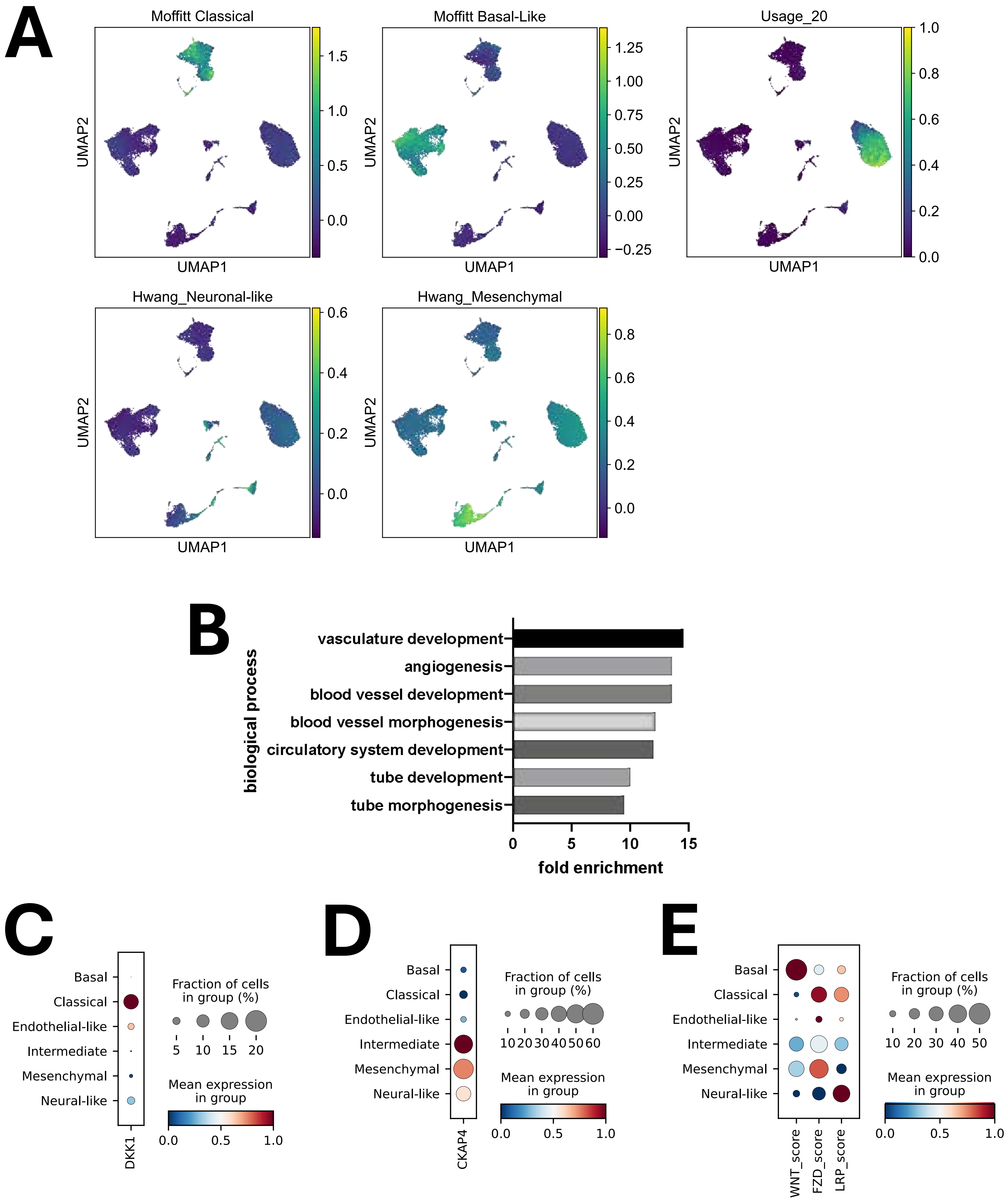
A
Moffitt Classical
Moffitt Basal-Like
Usage_20
UMAP2
UMAP1
1.5
1.0
0.5
0.0
1.25
1.00
0.75
0.50
0.25
0.00
−0.25
1.0
0.8
0.6
0.4
0.2
0.0
Hwang_Neuronal-like
Hwang_Mesenchymal
0.6
0.4
0.2
0.0
0.8
0.6
0.4
0.2
0.0
B
biological process
vasculature development
angiogenesis
blood vessel development
blood vessel morphogenesis
circulatory system development
tube development
tube morphogenesis
0
5
10
15
fold enrichment
C
Basal
Classical
Endothelial-like
Intermediate
Mesenchymal
Neural-like
DKK1
Fraction of cells in group (%)
5 10 15 20
Mean expression in group
0.0 0.5 1.0
D
Basal
Classical
Endothelial-like
Intermediate
Mesenchymal
Neural-like
CKAP4
Fraction of cells in group (%)
10 20 30 40 50 60
Mean expression in group
0.0 0.5 1.0
E
Basal
Classical
Endothelial-like
Intermediate
Mesenchymal
Neural-like
WNT_score
FZD_score
LRP_score
Fraction of cells in group (%)
10 20 30 40 50
Mean expression in group
0.0 0.5 1.0

Figure S10

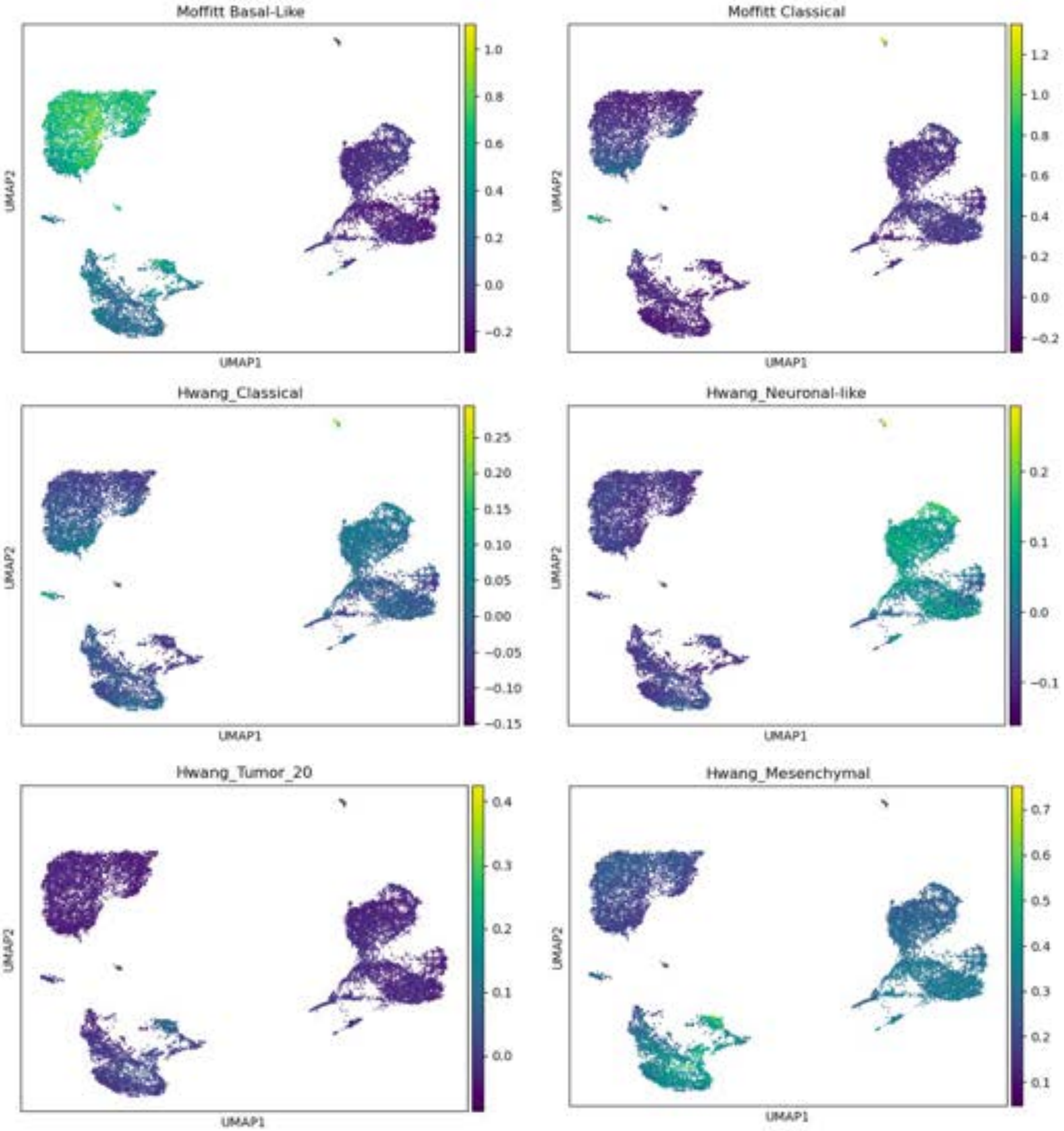
Moffitt Basal-Like
Moffitt Classical
Hwang_Classical
Hwang_Neuronal-like
Hwang_Tumor_20
Hwang_Mesenchymal
UMAP1
UMAP2

Figure S11

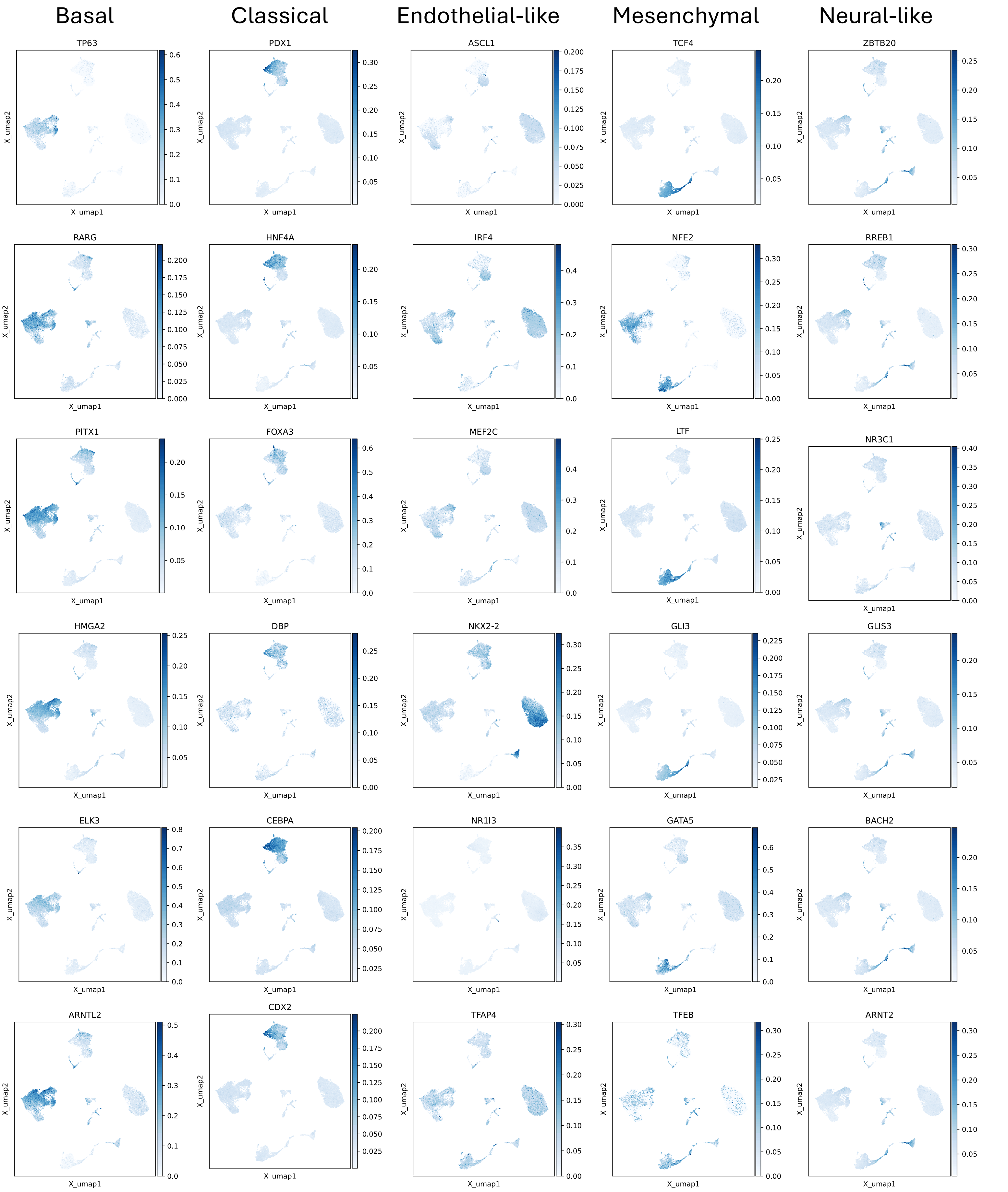
Basal
Classical
Endothelial-like
Mesenchymal
Neural-like
TP63
PDX1
ASCL1
TCF4
ZBTB20
RARG
HNF4A
IRF4
NFE2
RREB1
PITX1
FOXA3
MEF2C
LTF
NR3C1
HMGA2
DBP
NKX2-2
GLI3
GLIS3
ELK3
CEBPA
NR1I3
GATA5
BACH2
ARNTL2
CDX2
TFAP4
TFEB
ARNT2
X_umap1
X_umap2

Figure S12
